# Machine-learning-based prediction of disability progression in multiple sclerosis: an observational, international, multi-center study

**DOI:** 10.1101/2022.09.08.22279617

**Authors:** Edward De Brouwer, Thijs Becker, Lorin Werthen-Brabants, Pieter Dewulf, Dimitrios Iliadis, Cathérine Dekeyser, Guy Laureys, Bart Van Wijmeersch, Veronica Popescu, Tom Dhaene, Dirk Deschrijver, Willem Waegeman, Bernard De Baets, Michiel Stock, Dana Horakova, Francesco Patti, Guillermo Izquierdo, Sara Eichau, Marc Girard, Alexandre Prat, Alessandra Lugaresi, Pierre Grammond, Tomas Kalincik, Raed Alroughani, Francois Grand’Maison, Olga Skibina, Murat Terzi, Jeannette Lechner-Scott, Oliver Gerlach, Samia J. Khoury, Elisabetta Cartechini, Vincent Van Pesch, Maria Jose Sa, Bianca Weinstock-Guttman, Yolanda Blanco, Radek Ampapa, Daniele Spitaleri, Claudio Solaro, Davide Maimone, Aysun Soysal, Gerardo Iuliano, Riadh Gouider, Tamara Castillo-Triviño, Jose Luis Sanchez-Menoyo, Guy Laureys, Anneke van der Walt, Jiwon Oh, Eduardo Aguera-Morales, Ayse Altintas, Abdullah Al-Asmi, Koen de Gans, Yara Fragoso, Tunde Csepany, Suzanne Hodgkinson, Norma Deri, Talal Al-Harbi, Bruce Taylor, Orla Gray, Patrice Lalive, Csilla Rozsa, Chris McGuigan, Allan Kermode, Angel Perez sempere, Simu Mihaela, Magdolna Simo, Todd Hardy, Danny Decoo, Stella Hughes, Nikolaos Grigoriadis, Attila Sas, Norbert Vella, Yves Moreau, Liesbet Peeters

## Abstract

**Background:** Disability progression is a key milestone in the disease evolution of people with multiple sclerosis (PwMS). Prediction models of disability progression have not yet reached the level of trust needed to be adopted in the clinic. A common benchmark to assess model development in multiple sclerosis is also currently lacking.

**Methods:** Data of adult PwMS with a follow-up of at least three years from 146 MS centers, spread over 40 countries and collected by the MSBase consortium was used. With basic inclusion criteria for quality requirements, it represents a total of 15, 240 PwMS. External validation was performed and repeated five times to assess the significance of the results. TRIPOD guidelines were followed.

Confirmed disability progression after two years was predicted, with a confirmation window of six months. Only routinely collected variables were used such as the expended disability status scale, treatment, relapse information, and MS course.

To learn the probability of disability progression, state-of-the-art machine learning models were investigated. The discrimination performance of the models is evaluated on their area under the receiver operator curve (ROC-AUC) and under the precision recall curve (AUC-PR), and their calibration via the Brier score and the expected calibration error.

**Findings:** A temporal attention model was the best model. It achieved a ROC-AUC of 0·71 ± 0·01, an AUC-PR of 0·26 ± 0·02, a Brier score of 0·1 ± 0·01 and an expected calibration error of 0·07 ± 0·04. The history of disability progression is more predictive for future disability progression than the treatment or relapses.

**Interpretation:** Good discrimination and calibration performance on an external validation set is achieved, using only routinely collected variables. This makes these models ready for a clinical impact study. All our preprocessing and model code is available at https://gitlab.com/edebrouwer/ms_benchmark, making this task an ideal benchmark for predicting disability progression in MS.

## 1. Introduction

Multiple sclerosis (MS) is a chronic autoimmune disease of the central nervous system [1]. A recent census estimated more than 2·8 million people are currently living with MS [2]. It causes a wide variety of symptoms such as mobility problems, cognitive impairment, pain and fatigue. Importantly, the rate of disability progression is highly variable among people with MS (PwMS) [3]. This heterogeneity makes the personalization of care difficult and prognostic models are thus of high relevance for medical professionals, as they could contribute to better individualized treatment decisions. Indeed, a more aggressive treatment could be prescribed in case of a negative prognosis. Moreover, surveys indicate that PwMS are interested in their prognosis [4], which could help them with planning their lives.

There is a large amount of literature on prognostic MS models [5, 6, 7, 8, 9]. Some prognostic models are or were at some point available as web tools. However, with the exception of Tintore et al. [9] that focuses on conversion to MS, none have been integrated into clinical practice and no clinical impact studies have been performed [5, 6]. Because MS is a complex chronic disease that is often treated within a multidisciplinary context, the performance of a prognostic model studied in isolation from its clinical context gives limited information on its clinical relevance [10, 11]. Recent systematic reviews have highlighted several methodological issues within the current literature [5, 6], such as the lack of calibration or a possible significant bias in the cohort selection. Moreover, the investigated data sets are rarely made available. They furthermore often contain variables that are not routinely collected within the current clinical workflow (e.g. neurofilament light chain) or are not readily available for digital analysis (e.g. MRI).

In this article, we develop and externally validate a model to predict disability progression after two years for PwMS, using commonly-available clinical features.

This work was supported by a large project (Flanders AI), with all partners implementing different models. The best model was a temporal attention model with continuous temporal embedding.

Importantly, and in contrast with the available literature on disease progression models for MS (except for one model to predict relapses [12]), our data pre-processing pipeline and our models check all the boxes of the TRIPOD checklist. Our work therefore provides an important step towards the integration of artificial intelligence (AI) models in MS care.

## 2. Materials and Methods

### 2.1. Data source

In this multi-center international study, we used data of people with MS from 146 centers spread over 40 different countries and collected in the MSBase registry [13] as of September 2020. All data were prospectively collected during routine clinical care predominantly from tertiary MS centres [14]. The MSBase data set can be requested through MSBase that facilitates data sharing agreements with each individual center.

### 2.2. Inclusion criteria

The inclusion criteria for the initial extraction of the data from MSBase were at least 12 months of follow-up, aged 18 years or older, deceased or living and diagnosed with relapsing remitting (RR) MS, secondary progressive (SP) MS, primary progressive (PP) MS or clinically-isolated syndrome (CIS). This initial data set contained a total of 44,886 patients.

In order to ensure data quality, some patients were removed from that cohort. Exclusion criteria include:

- Visits of the same patient that happened on the same day but had different expanded disability status scale (EDSS) values were removed. All duplicate visits with the same EDSS for the same visit date were removed (i.e., only one of the visits was retained). Visits from before 1970 were discarded.
- Patients with the CIS MS course at their last visit were discarded. For those patients the relevant question is whether or not they will progress to confirmed MS, which is a different question than the one investigated in this work.

The complete list of exclusion criteria is available in Supplementary Section Appendix D. These criteria resulted in a total number of 40, 827 patients in the cohort. The details of the cohort construction are represented graphically in Figure 1.

**Figure 1:**
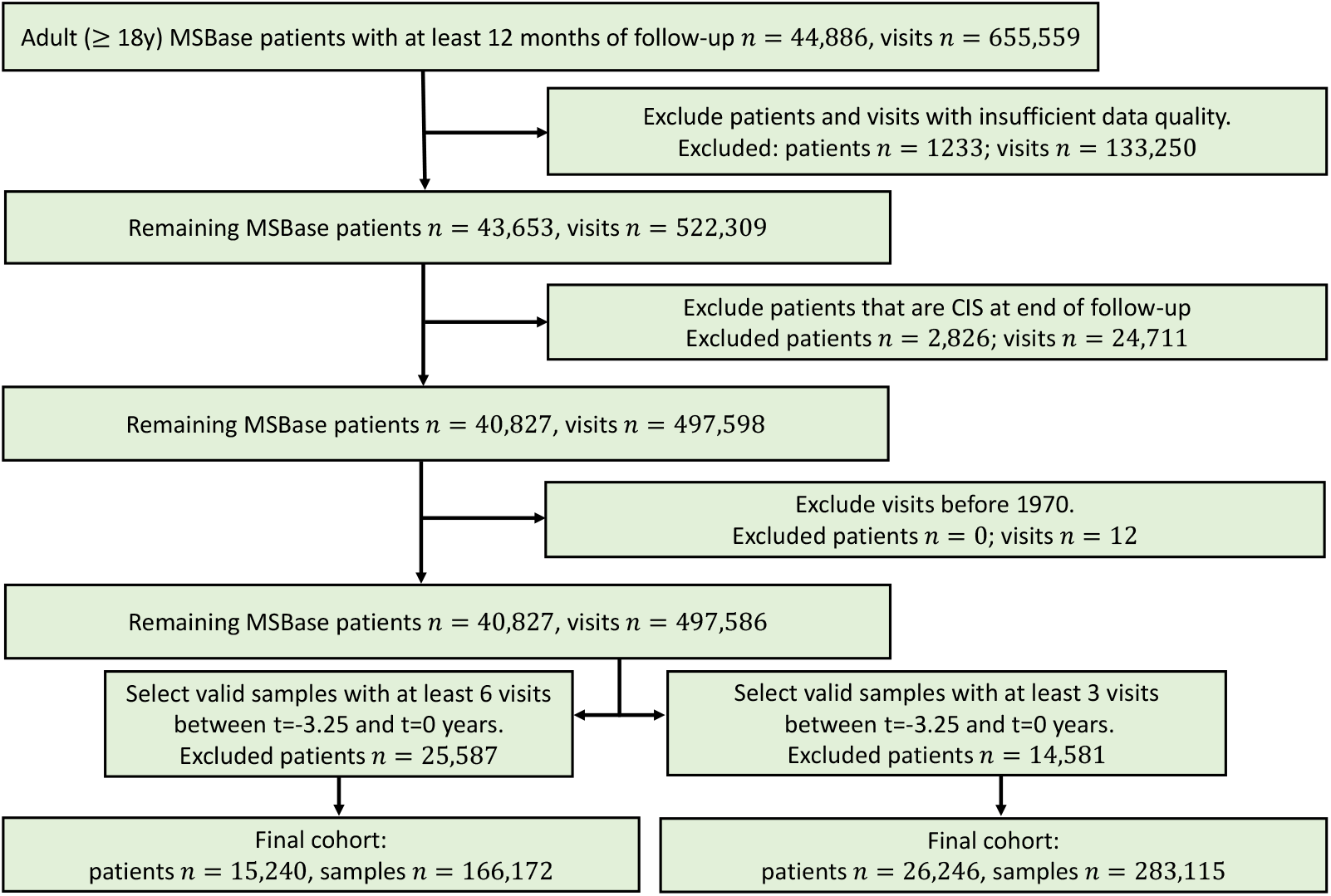
Flowchart of patient selection for both at least three and at least six visits in the last 3.25 years.

### 2.3. Definition of disability progression

Confirmed disability progression was defined based on the EDSS measurements. EDSS was scored by accredited scorers (Neurostatus certification was required at each center) and was calculated based on the functional system scores.

Because assessing progression requires a baseline EDSS value to compare with, predictions were made at visit dates where an EDSS measurement was recorded. In our notation, *t*_0_ denotes the time of the visits at which the prediction is made and the baseline EDSS is thus written as EDSS_*t*=0_. Motivated by the non-linearity of the EDSS, unconfirmed disability progression (*w* = 1)^1^ after two years (*t* = 2*y*) is defined as follows [15]:

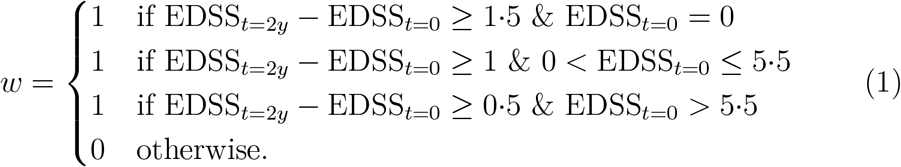

EDSS_*t*=2*y*_ represent the last recorded EDSS before *t* = 2 years.

EDSS suffers from inter- and intra-rater variability [16]. The actual state of the patient also fluctuates, because of e.g. recent relapses from which the patient could still (partly) recover. We therefore study *confirmed* disability progression (*w*_*c*_) for at least six months. Progression is confirmed if all EDSS values measured within six months after the progression event and the first EDSS measurement after two years lead to the same worsening target *w*_*u*_ = 1 according to Eq. 1. EDSS measurements within one month after a relapse are not taken into account for confirming disability progression [15].

### 2.4. Definition of clinical episodes

For each patient, all visits can potentially represent a valid EDSS baseline for a progression episode. More generally, it is possible to divide the available clinical history of a patient in multiple (potentially overlapping) episodes for which a disability progression label can be computed. Each episode therefore consists of an observation window, a baseline EDSS (EDSS_*t*=0_) and a confirmation label (*w*_*c*_) as shown on Figure 2. Extracting several episodes per patient allows to significantly increase the number of data points in the study.

**Figure 2:**
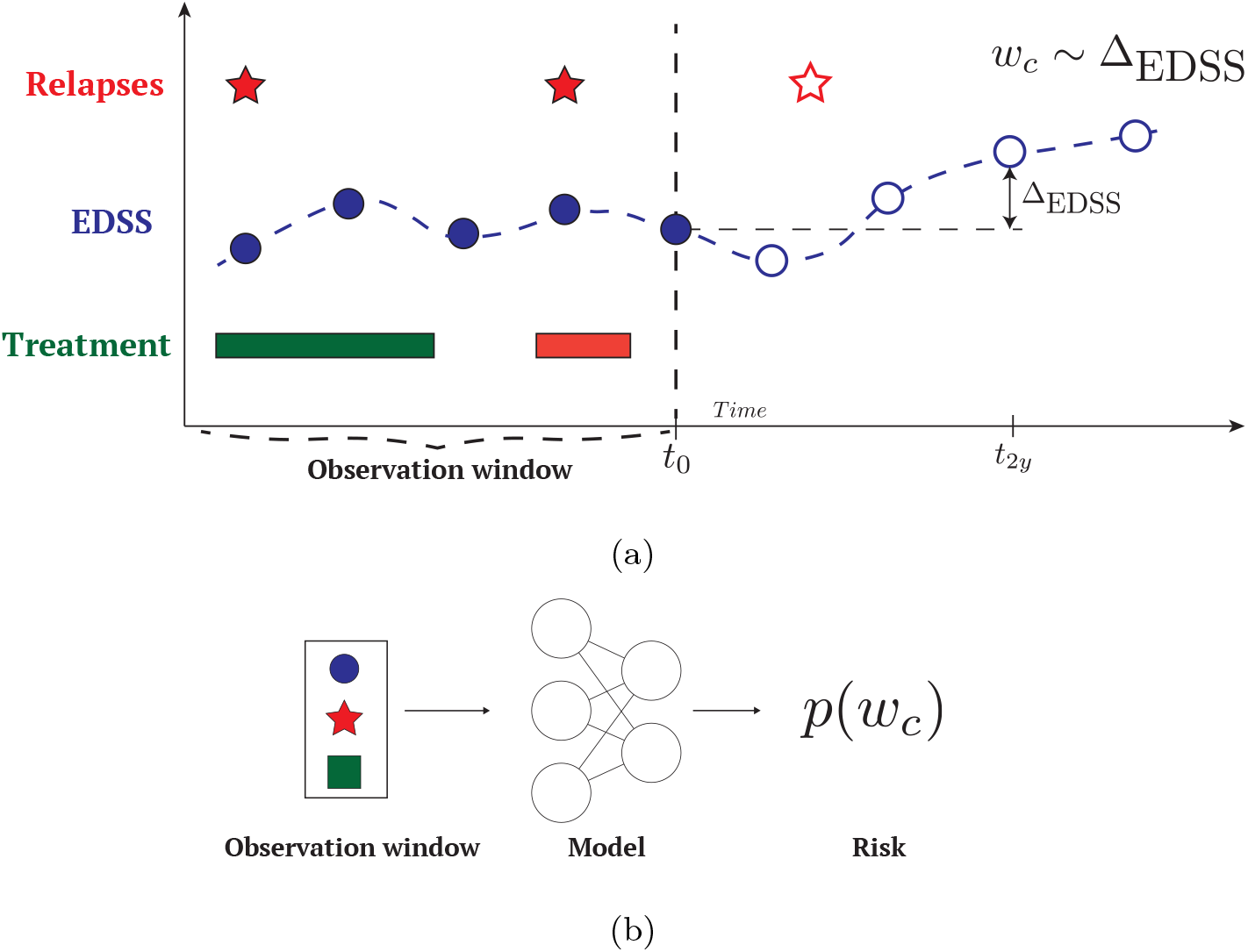
Problem Setup. (a) For each patient episode, the available data for prediction consists of the baseline data and the longitudinal clinical data in the observation window. Disability progression (*w*_*c*_) is assessed based on the difference between the EDSS at time *t* = 0 and two years later 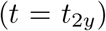 as defined in Equation 1. (b) Based on the available historical clinical data (in the observation time window), we aim at training a model able to predict the risk *p*(*w*_*c*_) of disability progression at a two years horizon 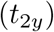.

We define two cohorts of patients, one with a minimum of three EDSS measurements, the other with a minimum of six EDSS measurements over the last three years and three months of the observation window. The final cohorts resulted in a total of 283,115 valid episodes from 26,246 patients, for a minimum of three EDSS measurements and 166,172 valid episodes from 15,240 patients, for a minimum of six EDSS measurements.

### 2.5. Variables

A set of clinical variables was retained from all available variables and included in the observation window of each episode. The following static (*i.e*., non-varying over time) variables were selected: birth date, sex, MS onset date, education status (higher education, no higher education, unknown) and the location of the first symptom (*i.e*., supratentorial, optic pathways, brainstem or spinal cord).

The following longitudinal variables were also collected in the observation window (*i.e*., for times *t* ≤ 0): EDSS, MS course (RRMS, PPMS, SPMPS, CIS), relapse occurrence, relapse position (pyramidal tract, brainstem, bowel bladder, cerebellum, visual function, sensory), all Kurtzke functional system (KFS) scores, Fampridine administration. The disease modifying therapies (DMT) and immunosuppressants were categorized into low-efficacy, moderate-efficacy and high-efficacy:

- Low-efficacy: Interferons, Teriflunomide, Glatiramer, Azathioprine, Methotrexate.
- Moderate-efficacy: Fingolimod, Dimethyl-Fumarate, Cladribine, Siponimod, Daclizumab
- High-efficacy: Alemtuzumab, Rituximab, Ocrelizumab, Natalizumab, Mitoxantrone, Cyclophosphamide.

Except for Mitoxantrone and Cyclophosphamide, we assumed that only one DMT was administered at the same time.

MRI variables were not included due to high missingness. Indeed, the lesion counts were available in less than 1·7% of the clinical episodes. The variable indicating whether the MRI was normal, abnormal MS typical, or abnormal MS atypical was judged as not informative enough.

The above variables were then grouped in three feature sets: *static, dynamic* (summary statistics of the clinical history) and *longitudinal* [17]. These represent increasing quantity of information regarding the clinical history of patients. More details regarding the variables used and the grouping can be found in Supplementary Section Appendix H.

### 2.6. Models

The disability progression was framed as a classification problem. The following models were used to predict disability progression: a temporal attention model with continuous temporal embeddings [18], a Bayesian neural network and a multi-layer perceptron. This work was supported by a large project (Flanders AI) and those models were selected as the best performing ones among a larger array of candidate models implemented by the different partners (see Supplementary Section Appendix G for details). We follow the TRIPOD guidelines for reporting prognostic models [19]. The checklist can be found in Supplementary Figure E.8.

The multi-layer perceptron model is a neural network architecture that takes as input the static and dynamic features set, represented as a fixed length vector. The model is composed of five hidden layers of dimension 128.

The Bayesian neural network has a similar architecture as the multi-layer perceptron, but provides uncertainty estimates on the weights of the last hidden layer by incorporating MCdropout [20]. This should confer better generalization capabilities as well as better calibration.

The temporal attention model relies on a transformer architecture [18]. In contrast to the previous models, this architecture is able to handle the longitudinal feature set, as it is able to process the whole clinical time series. Each visit is encoded as a fixed-length vector along with a mask for missing features and a continuous temporal embedding. This temporal embedding allows for arbitrary time differences between measurements, and is therefore especially suited for clinical time series where irregular sampling is most common. The static and the dynamic feature sets were included in the model as extra temporal features that are repeated over the patient history. Two temporal attention layers with dimension 128 were used.

### 2.7. Evaluation

The data set was split into 60% for training, 20% for validation and 20% for testing. The validation data was used to optimize the hyperparameters of the models. Post-hoc calibration methods (Platt scaling [21] and isotonic regression [22]) were used on the validation set and the performance evaluated on the test set. The test set was not seen during model training and hyperparameter optimization. To produce a measure of uncertainty of the performance of the models, the procedure of splitting the data and training the models was repeated five times, corresponding to five splits.

As the data set consists of patients from different centers, we split the data set such that the validation and test sets represent an external validation. Patients from the same centers were therefore assigned to the same set (training, validation or test).

Discrimination was evaluated using the area under the receiving operator characteristic (ROC-AUC) and the area under the precision recall curve (AUC-PR). Calibration was evaluated numerically using the Brier score and the expected calibration error (ECE) with 20 bins. Calibration was evaluated visually using reliability diagrams.

## 3. Results

### 3.1. Participants

A flowchart of the patient inclusion for the final cohort is shown in Figure 1. The requirements on data quality and availability led to a final cohort of 15,240 and 25,246 for the three and six EDSS measurements criteria respectively. Basic characteristics of the final cohorts are shown in Table 1.

**Table 1:** Summary statistics of the cohort of interest after extraction from MSBase (Extracted Cohort) and after patient and sample selection (Final Cohort). For all variables the value at the last recorded visit was used.

| Variable | Cohort 3 EDSS | Cohort 6 EDSS |
| --- | --- | --- |
| Patients (% female) | 26,246 (71.8) | 15,240 (72.0) |
| Age, Years<sup>a</sup> | 42.8 (10.8) | 41.9 (10.1) |
| Age at MS onset, years<sup>a</sup> | 31.3 (8.9) | 30.6 (8.6) |
| Disease duration, years<sup>a</sup> | 11.6 (8.0) | 11.4 (7.3) |
| Education status, % higher<sup>c</sup> | 18.2 (65.1) | 16.6 (66.2) |
| First symptom, none given (%) | 13.7 | 13.0 |
| supratentorial (%) | 28.2 | 30.6 |
| optic pathways (%) | 22.6 | 23.2 |
| brainstem (%) | 24.3 | 25.7 |
| spinal cord (%) | 26.4 | 25.8 |
| MS course | / | / |
| CIS (%) | 0 | 0 |
| Relapsing-Remitting (%) | 83.5 | 85.7 |
| Primary Progressive (%) | 5.0 | 4.1 |
| Secondary Progressive (%) | 11.5 | 10.2 |
| EDSS<sup>a</sup> | 3.0 (2.1) | 3.1 (2.0) |
| Annualized relapse rate<sup>b</sup> | 0.82 [0.43, 1.47] | 0.86 [0.49, 1.43] |
| KFS Scores | / | / |
| pyramidal<sup>b</sup> | 2 [1, 3] | 2 [1, 3] |
| cerebellar<sup>b</sup> | 0 [0, 2] | 0 [0, 2] |
| brainstem<sup>b</sup> | 0 [0, 1] | 0 [0, 1] |
| sensory<sup>b</sup> | 1 [0, 2] | 1 [0, 2] |
| sphincteric<sup>b</sup> | 0 [0, 1] | 0 [0, 1] |
| visual<sup>b</sup> | 0 [0, 1] | 0 [0, 1] |
| cerebral<sup>b</sup> | 0 [0, 1] | 0 [0, 1] |
| ambulatory<sup>b</sup> | 0 [0, 1] | 0 [0, 1] |
| DMT | / | / |
| none | 23.5 | 19.0 |
| low-efficacy | 51.3 | 52.0 |
| moderate-efficacy | 13.6 | 14.8 |
| high-efficacy | 11.6 | 14.1 |
| high induction | 7.2 | 7.4 |

*a*: mean ± standard deviation
*b*: median (quartiles)
*c*: % missing data

### 3.2. Model development

The inclusion criteria and pre-processing of the raw data resulted in 283,115 episodes from 26,426 patients in the 3-visits cohort. 11·64% of those episodes represent a progression event, hence showing a mild imbalance. We addressed this imbalance by re-weighing each sample proportionally to its label occurrence. The code for training the models and the final models are publicly available and can be found at https://gitlab.com/edebrouwer/ms_benchmark.

### 3.3. Model performance

The performance of the models is reported in Tables 2 to 4. A visual illustration of the discrimination performance is shown in Figure 3. The attention-based model reaches a ROC-AUC of 0·71 ± 0·01 and a AUC under the precision-recall curve of 0·26 ± 0·02 with a calibration error of 0·07 ± 0·04 on the external test cohort.

**Table 2:** Summary statistics of the performance measures. Baseline performance are 0·5 for ROC-AUC and 0·11 for *AUC* − *PR*. ↑ indicates higher is better. ↓ indicates lower is better.

| Model | ROC-AUC ↑ | AUC-PR ↑ | Brier ↓ | ECE ↓ |
| --- | --- | --- | --- | --- |
| Ensemble | $0.71 \pm 0.01$ | $0.25 \pm 0.02$ | $0.10 \pm 0.01$ | $0.06 \pm 0.05$ |
| Attention | $0.71 \pm 0.01$ | $0.26 \pm 0.02$ | $0.10 \pm 0.01$ | $0.07 \pm 0.04$ |
| Bayesian NN | $0.71 \pm 0.01$ | $0.25 \pm 0.01$ | $0.10 \pm 0.01$ | $0.08 \pm 0.04$ |
| MLP | $0.70 \pm 0.01$ | $0.24 \pm 0.02$ | $0.10 \pm 0.01$ | $0.09 \pm 0.03$ |

**Figure 3:**
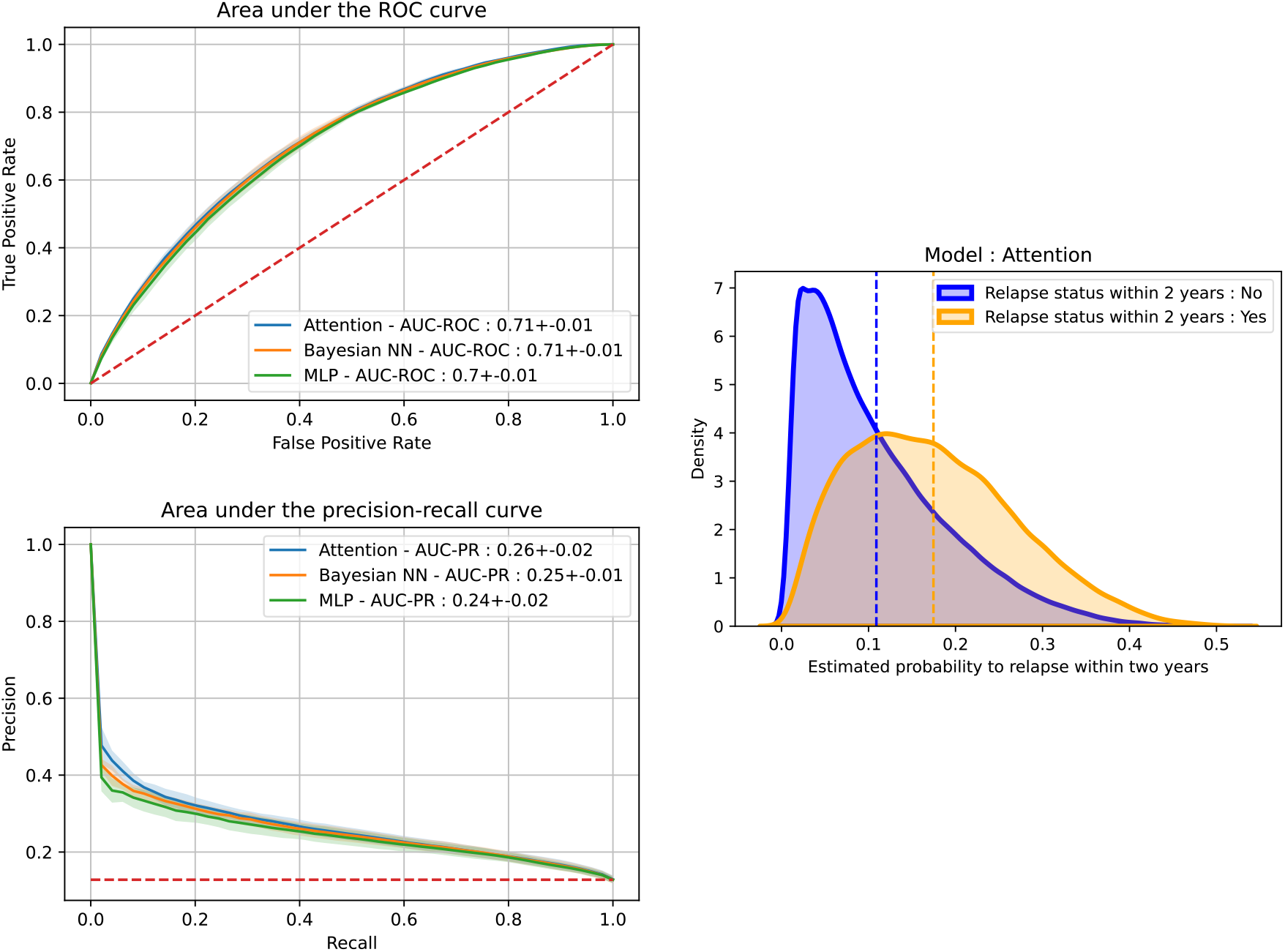
Visual representation of the discrimination performance: the ROC-AUC curve, the AUC-PR curve and the distribution of the estimated probability of relapse per group obtained with the temporal transformer model.

To assess the reliability of those results on specific sub-groups of patients, we also report the performance for each different MS course at time of prediction (Table 3) and different base EDSS (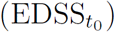) (Table 4). The relapsing-remitting (RR) category shows a performance similar to the full cohort. The smaller primary progressive and secondary progressive groups, on the other hand, suffer from low sample size, resulting in a decreased discrimination performance. The same effect is to be observed when we segment the performance by disability severity, with the group of higher severity showing a lower discrimination performance. In the supplementary material, we also show a segmentation of the results by the medical center of origin of the patients (Figure A.5), indicating a higher variability of the results for small centers.

**Table 3:** Results for disability progression prediction per MSCourse, for the best models. ↑ indicates higher is better. ↓ indicates lower is better.

| Model | MSCourse | ROC-AUC ↑ | AUC-PR ↑ | Brier ↓ | ECE ↓ |
| --- | --- | --- | --- | --- | --- |
| Attention | PP | $0.65 \pm 0.01$ | $0.33 \pm 0.04$ | $0.16 \pm 0.01$ | $0.07 \pm 0.02$ |
| Attention | RR | $0.70 \pm 0.01$ | $0.21 \pm 0.01$ | $0.09 \pm 0.01$ | $0.06 \pm 0.03$ |
| Attention | SP | $0.65 \pm 0.01$ | $0.33 \pm 0.03$ | $0.17 \pm 0.01$ | $0.10 \pm 0.05$ |
| Bayesian NN | PP | $0.66 \pm 0.01$ | $0.34 \pm 0.03$ | $0.16 \pm 0.01$ | $0.09 \pm 0.05$ |
| Bayesian NN | RR | $0.70 \pm 0.01$ | $0.20 \pm 0.01$ | $0.09 \pm 0.01$ | $0.09 \pm 0.03$ |
| Bayesian NN | SP | $0.64 \pm 0.01$ | $0.32 \pm 0.02$ | $0.17 \pm 0.01$ | $0.11 \pm 0.02$ |
| MLP | PP | $0.63 \pm 0.03$ | $0.32 \pm 0.05$ | $0.16 \pm 0.01$ | $0.09 \pm 0.03$ |
| MLP | RR | $0.69 \pm 0.01$ | $0.19 \pm 0.0$ | $0.09 \pm 0.01$ | $0.05 \pm 0.02$ |
| MLP | SP | $0.63 \pm 0.01$ | $0.31 \pm 0.02$ | $0.17 \pm 0.01$ | $0.10 \pm 0.04$ |

**Table 4:**
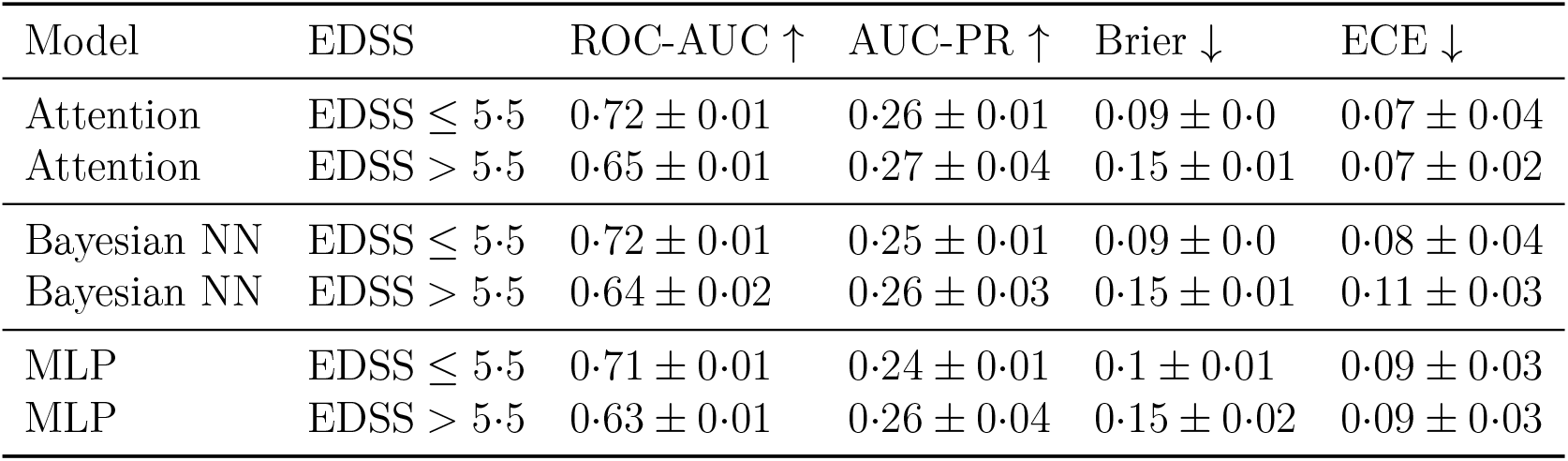
Results for disability progression prediction for EDSS ≤ 5·5 and *>* 5·5. ↑ indicates higher is better. ↓ indicates lower is better.

The calibration of the different models can be assessed from the Brier score and the expected calibration errors from the results tables. In Figure 4, we also show the calibration plot of the longitudinal attention model on the external test cohort. We observe a very good calibration of the predicted risks in the range between 0 and 0.3, suggesting an excellent reliability of the predictive model. The calibration curves of other models are given in the supplementary materials (Figure B.6) along with a segmentation of the calibration of the models by clinical subgroups (Figure C.7).

**Figure 4:**
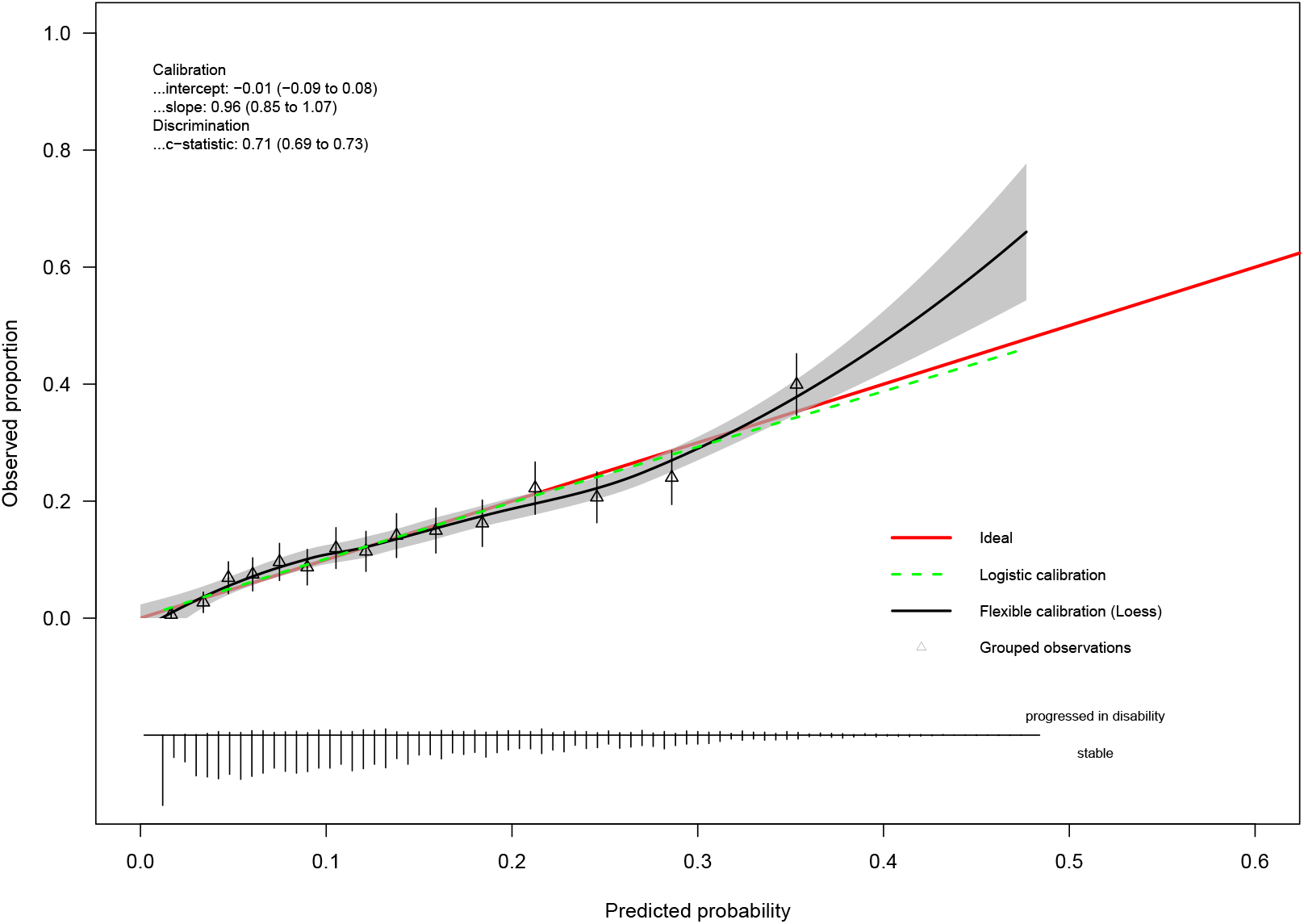
Calibration diagram for the Attention model for the first data split. The *val.prob.ci.2* function [23] was used.

### 3.4. Feature importance

The importance of the different variables used in our models is investigated. Table 5 shows the results of a permutation importance test on the MLP model, by assessing the loss in discrimination performance when a variable is shuffled over the test set [24]. Table 5 ranks the features in decreasing order of importance. The most important variables include the baseline EDSS at prediction time, the number of years since 1990 and the mean EDSS and KFS over the last 3 years.

**Table 5:**
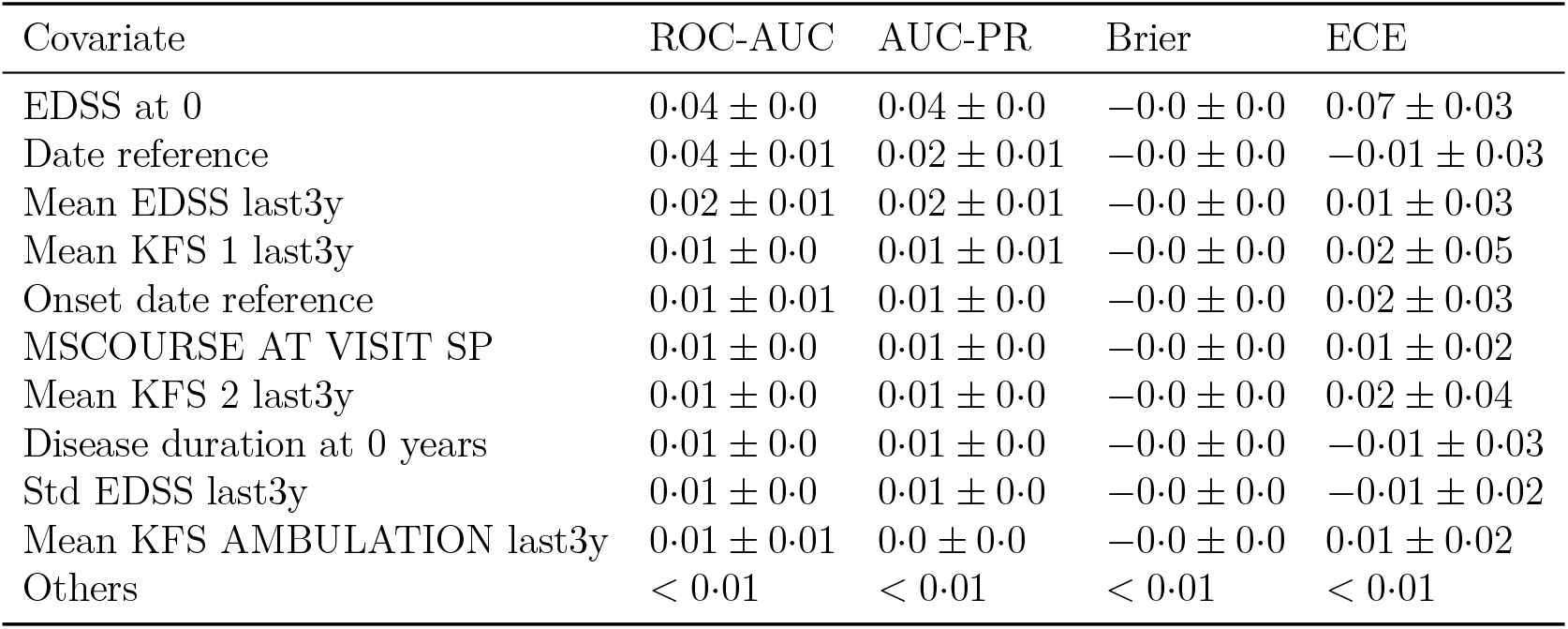
Features ranked by order of importance for the Dynamic Model. Feature importance is assessed by the average difference in ROC-AUC when the specific feature is shuffled.

| Covariate | ROC-AUC | AUC-PR | Brier | ECE |
| --- | --- | --- | --- | --- |
| EDSS at 0 | $0.04 \pm 0.0$ | $0.04 \pm 0.0$ | $-0.0 \pm 0.0$ | $0.07 \pm 0.03$ |
| Date reference | $0.04 \pm 0.01$ | $0.02 \pm 0.01$ | $-0.0 \pm 0.0$ | $-0.01 \pm 0.03$ |
| Mean EDSS last3y | $0.02 \pm 0.01$ | $0.02 \pm 0.01$ | $-0.0 \pm 0.0$ | $0.01 \pm 0.03$ |
| Mean KFS 1 last3y | $0.01 \pm 0.0$ | $0.01 \pm 0.01$ | $-0.0 \pm 0.0$ | $0.02 \pm 0.05$ |
| Onset date reference | $0.01 \pm 0.01$ | $0.01 \pm 0.0$ | $-0.0 \pm 0.0$ | $0.02 \pm 0.03$ |
| MSCOURSE AT VISIT SP | $0.01 \pm 0.0$ | $0.01 \pm 0.0$ | $-0.0 \pm 0.0$ | $0.01 \pm 0.02$ |
| Mean KFS 2 last3y | $0.01 \pm 0.0$ | $0.01 \pm 0.0$ | $-0.0 \pm 0.0$ | $0.02 \pm 0.04$ |
| Disease duration at 0 years | $0.01 \pm 0.0$ | $0.01 \pm 0.0$ | $-0.0 \pm 0.0$ | $-0.01 \pm 0.03$ |
| Std EDSS last3y | $0.01 \pm 0.0$ | $0.01 \pm 0.0$ | $-0.0 \pm 0.0$ | $-0.01 \pm 0.02$ |
| Mean KFS AMBULATION last3y | $0.01 \pm 0.01$ | $0.0 \pm 0.0$ | $-0.0 \pm 0.0$ | $0.01 \pm 0.02$ |
| Others | $< 0.01$ | $< 0.01$ | $< 0.01$ | $< 0.01$ |

The baseline EDSS is expected to be important in the prediction, as the definition of the progression event directly depends on it (as seen in Eq. 1). The time since 1990 suggests a change of behavior of the disease over the years that could be explained by progress in clinical care or enhanced diagnosis of earlier and milder forms of the disease. Remarkably, the importance of the previous values of EDSS and KFS demonstrates an added value of considering longitudinal data, as already shown in Brouwer et al. [17]. Remarkably, no variables including DMTs are given any significant importance.

## 4. Discussion

The models investigated in this study provide a significant advance towards deploying AI in clinical practice in MS. After validation of the results in a clinical impact study, they have the potential to let the MS field benefit from the advantages of advanced predictive modelling capabilities.

Our work confirms that predicting disability progression of MS patients is feasible. Importantly, it can be achieved with variables that are collected as part of routine clinical care. Despite MS progression being inherently stochastic, we show that relevant historical clinical data can lead to high discrimination performance combined with a good calibration (Figure 4), which is crucial in healthcare applications. This points towards a readiness of this model to be tested in a clinical impact study. Our external validation procedure ensures generalizability within centers participating in MSBase, as it allows to estimate inter-center variation of the performance.

However, the models also suffer from limitations. First of all, several countries with good quality MS registries were not included because they are not part of the MSBase initiative. Since treatment decisions can be country specific to a significant degree [25], it can result in a difference of performance of the proposed models on countries not included in this data set. Yet, a clinical impact study in MS centers participating in MS Base would not suffer from such external validity problems.

Second, our inclusion criteria require patients with good follow-up (at least one yearly visit with EDSS measurement), so stable patients that do not visit regularly cannot benefit from this model. Importantly, because we rely on at least three years of historical clinical data, our model cannot be applied for patients who got recently diagnosed with MS. This task for newly-diagnosed patients would require the design of a dedicated model.

Third, the progression target that we defined in this work cannot realistically fully capture the complexity of the disease and progression in MS cannot be summarized by EDSS only. Despite its imperfections, this metric has been proven clinically useful, striking a good balance between abstraction and expressiveness. Our work therefore builds upon those concepts and inherits their flaws and advantages.

Despite these imperfections, our models could potentially help patients in the planning of their lives and provide a baseline for further research. An emphasis on reproducibility was made, in an attempt to provide a strong benchmark for this important task. Thanks to the excellent clinically-informed pre-processing pipeline, researchers can easily extend the current models or propose their own, to continuously improve disease progression prediction. Extensions to our method could include treatment recommendation or inclusion of other biomarkers available in a specific center.

## 5. Conclusion

In this work, we developed and externally validated machine learning models for predicting disability progression of people with MS. The performance achieved by these models, along with the availability of the predictors they rely on, implies that a clinical impact study is feasible. Such an impact study would provide important information regarding how patients use such model predictions, and how medical professionals interact and use such predictions.

Our clinically-informed data processing pipeline and task definition allow the machine learning community to contribute meaningfully in improving such prediction models.

### Data Sharing

The data set used in this study is available upon request to the MSBase principal investigators included in the study. MSBase operates as a single point of contact to facilitate the data sharing agreements with the individual data custodians.

## Data Availability

The data used in study originates from the MSBase data set which can be requested through the MSBase organization that facilitates data sharing agreements with each individual center.

https://www.msbase.org/

## Acknowledgements

Edward De Brouwer is funded by a FWO-SB grant. This research received funding from the Flemish Government under the “Onderzoeksprogramma Artificiële Intelligentie (AI) Vlaanderen” programme.

## Appendix A. Per-center validation

On Figure A.5, we plot the ROC-AUC of individual centers in the test set against the size of the center. We observe that as the size of the centers grow, the performance converges to the average ROC-AUC. As the size of centers shrinks, the variability in performance increases, which is statistically expected due to low sampled size.

**Figure A.5:**
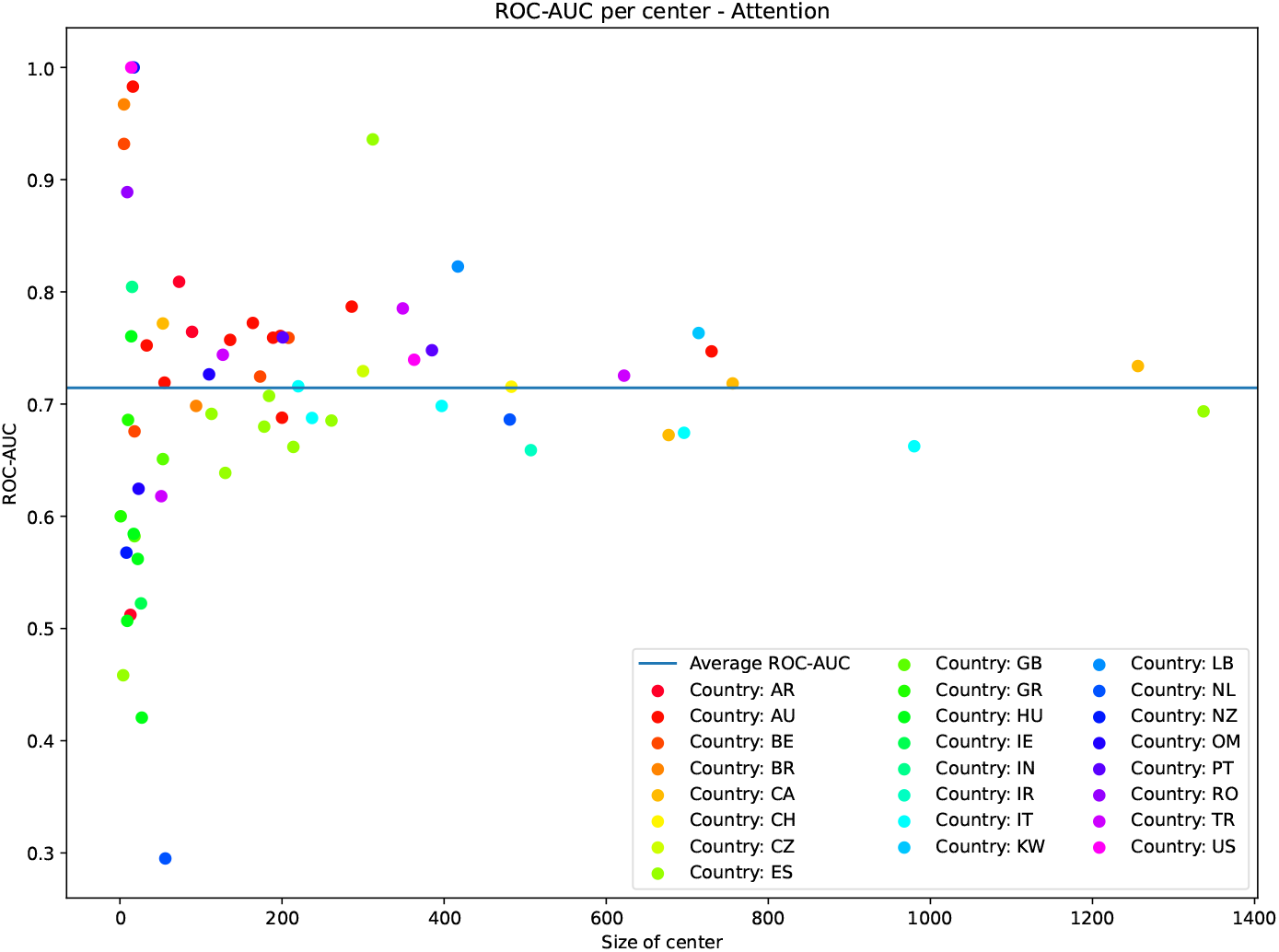
AUROC scores per center, example from paper. Centers with no progression are not plotted (because AUC not defined)

## Appendix B. Calibration curves

On Figure B.6, we show the calibration curves of the different models on the test set (fold (*e.g*. train-test split) 0). Calibration was performed using Platt scaling [21]. We observe good calibration for all models. The discrepancy with the ideal line (dotted) in the larger scores regime can be explained by the lowest number of data points in that region, leading to more variance.

**Figure B.6:**
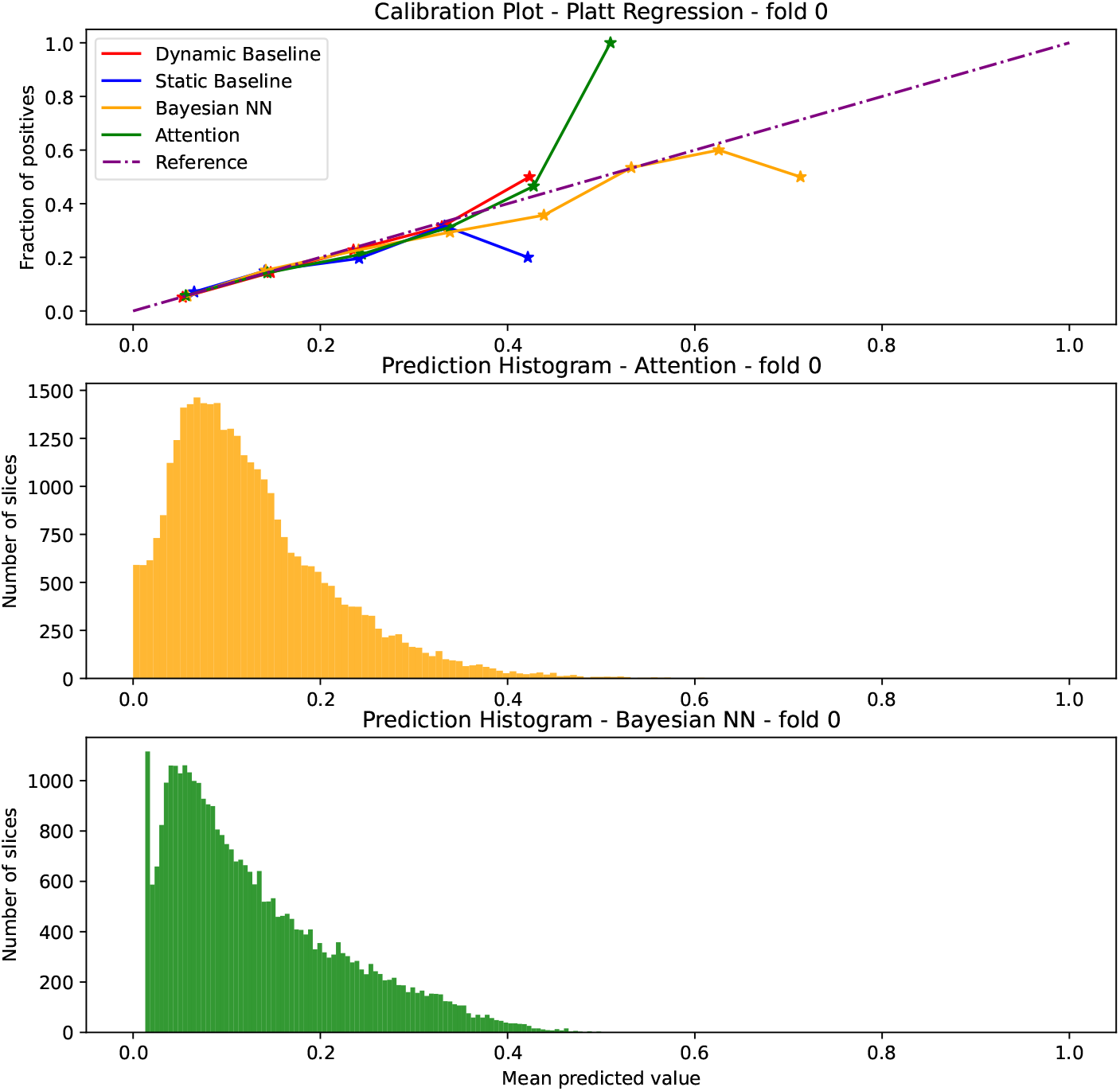
Calibration diagram for all models. For the 1st data split.

## Appendix C. Calibration per clinical group

In Figure C.7, we show the prevalence of progression within different clinical subgroups of patients (Observed proportion) and the average probability of progression in the subgroup as given by the different models. We observe an acceptable discrepancy (of maximum 3 points), and a tendency of the models to underestimate the prevalence of disability progression.

**Figure C.7:**
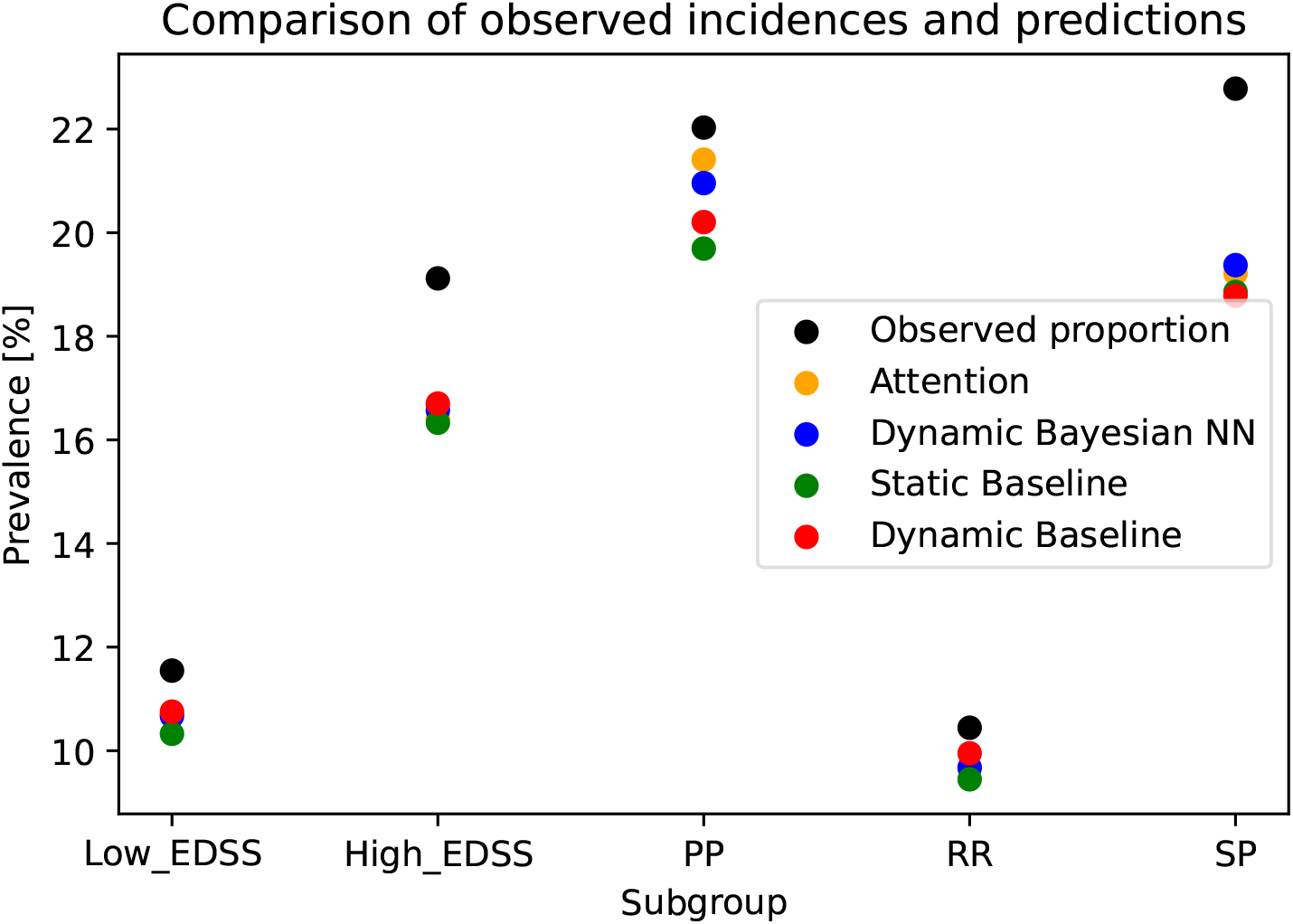
Predicted percentage of worsening per subgroup, for both MS Courses and EDSS larger or smaller than 5.5. (In this figure green is the actual prevalence for the age groups on the x-axis, and red and purple are model predictions. For us this would be MS course and high or low EDSS). This is a way to assess calibration performance for different subgroups.

## Appendix D. Exclusion criteria

- Patients whose diagnosis date or age at first symptoms (i.e., MS onset date) was missing or with invalid formatting were removed.
- Patient whose MS course or sex was not available were removed.
- Patients whose date of MS diagnosis, birth, MS onset, start of progression, clinic entry or first relapse was higher than the extraction date were discarded.
- All visits whose visit date had an invalid format or was after the extraction date were discarded.
- Visits of the same patient that happened on the same day but had different expanded disability status scale (EDSS) values were removed. All duplicate visits with the same EDSS for the same visit date were removed (i.e., only one of the visits was retained). Visits from before 1970 were discarded.
- Patients with the CIS MS course at their last visit were discarded. For those patients the relevant question is whether or not they will progress to confirmed MS, which is a different question than the one investigated in this work.

## Appendix E. Tripod checklist

The design of the algorithms carefully followed the TRIPOD checklist as shown on Figure E.8. All points are checked or are deemed not applicable in our study. This consists of the following :

- 6b. Report any actions to blind assessment of the outcome to be predicted.
- 11. *Provide details on how risk groups were created, if done*. No risk groups were identified in this study.
- 14b. This can only be done for statistical models. However, we report measures of variables importance in section 3.4.
- 17. *Model updating*. The models proposed here are not updates of previous iterations but rather their first development.

Note also that no sample size calculations were performed; the size of this retrospective data set was fixed.

## Appendix F. Full comparison with other machine-learning models

In this section, we report more detailed performance results of the proposed models along with other machine-learning architecture that were considered. More information about all considered architectures is to be found in Appendix G.

**Figure E.8:**
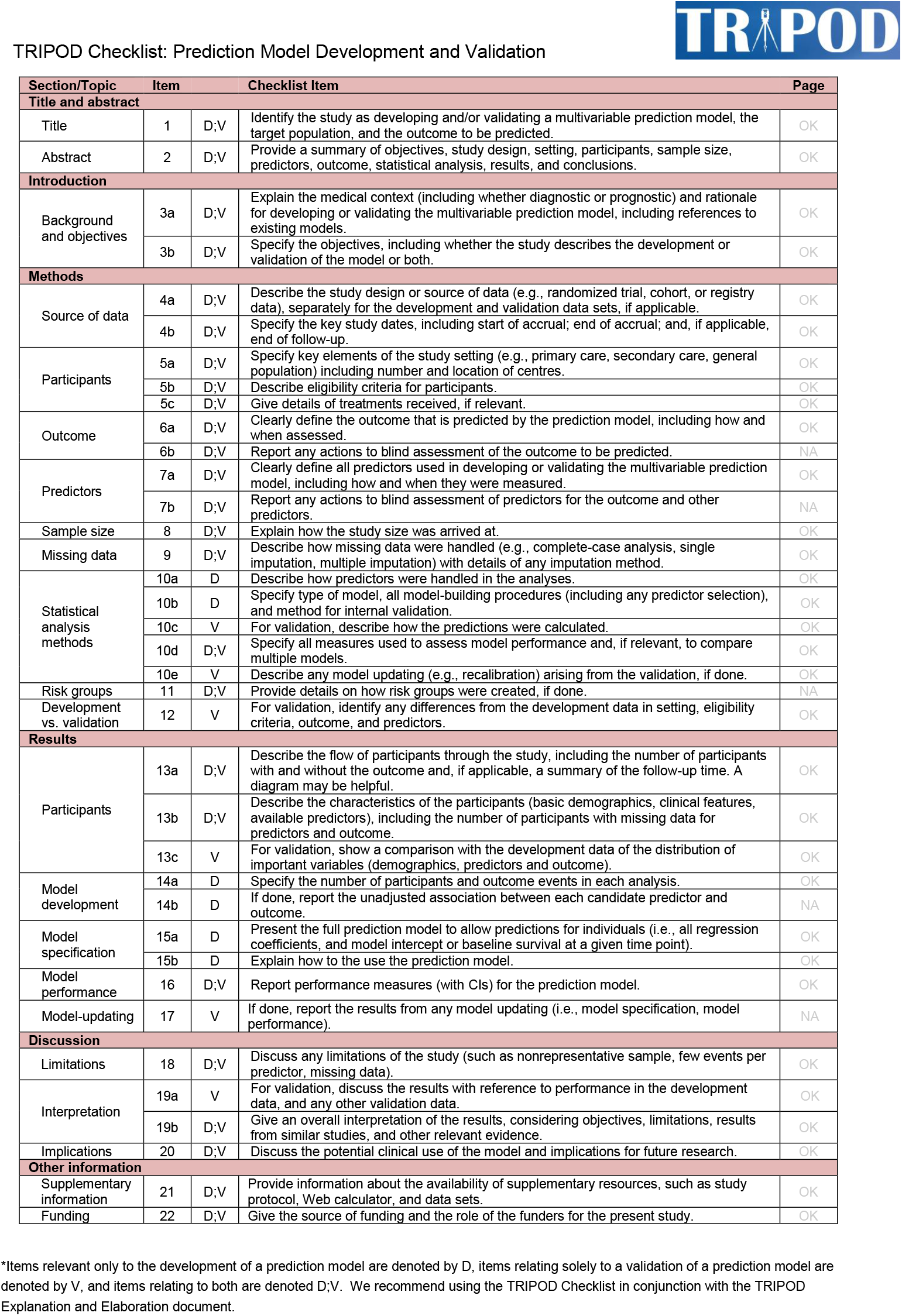
TRIPOD checklist

### Appendix F.1. Overall performance

Tables F.6 and F.7 report the ROC-AUC, AUC-PR, Brier Score and ECE of all models. Two cohorts are considered: patients with a least 3 visits with EDSS in the last 3.25 years and patients with at least 6 visits with EDSS in the last 3.25 years.

**Table F.6:** Summary statistics of the performance measures. Cohort with minimum 3 visits.

| Model | ROC-AUC | AUC-PR | Brier | ECE |
| --- | --- | --- | --- | --- |
| Ensemble | $0.71 \pm 0.01$ | $0.25 \pm 0.02$ | $0.10 \pm 0.01$ | $0.06 \pm 0.05$ |
| Attention | $0.71 \pm 0.01$ | $0.26 \pm 0.02$ | $0.10 \pm 0.01$ | $0.07 \pm 0.04$ |
| RNN | $0.71 \pm 0.01$ | $0.25 \pm 0.02$ | $0.10 \pm 0.01$ | $0.04 \pm 0.02$ |
| Static Bayesian NN | $0.67 \pm 0.01$ | $0.22 \pm 0.02$ | $0.11 \pm 0.01$ | $0.09 \pm 0.05$ |
| Dynamic Bayesian NN | $0.71 \pm 0.01$ | $0.25 \pm 0.01$ | $0.10 \pm 0.01$ | $0.08 \pm 0.04$ |
| Static Baseline | $0.67 \pm 0.01$ | $0.22 \pm 0.02$ | $0.11 \pm 0.01$ | $0.09 \pm 0.05$ |
| Dynamic Baseline | $0.70 \pm 0.01$ | $0.24 \pm 0.02$ | $0.10 \pm 0.01$ | $0.09 \pm 0.03$ |
| Static Logistic | $0.66 \pm 0.02$ | $0.21 \pm 0.02$ | $0.11 \pm 0.01$ | $0.04 \pm 0.01$ |
| Dynamic Logistic | $0.7 \pm 0.01$ | $0.24 \pm 0.02$ | $0.11 \pm 0.01$ | $0.06 \pm 0.02$ |
| Static DeepMTP | $0.66 \pm 0.03$ | $0.21 \pm 0.03$ | $0.11 \pm 0.01$ | $0.23 \pm 0.06$ |
| Dynamic DeepMTP | $0.68 \pm 0.02$ | $0.23 \pm 0.02$ | $0.11 \pm 0.01$ | $0.17 \pm 0.04$ |
| Static FactorizationMachine | $0.67 \pm 0.01$ | $0.22 \pm 0.02$ | $0.11 \pm 0.01$ | $0.18 \pm 0.07$ |
| Dynamic FactorizationMachine | $0.69 \pm 0.01$ | $0.24 \pm 0.02$ | $0.11 \pm 0.01$ | $0.17 \pm 0.02$ |

### Appendix F.2. Performance per MS course

Tables F.8 and F.9 report the the ROC-AUC, AUC-PR, Brier Score and ECE of all models on the different MS course subgroups. Primary Progressive (PP), Relapsing Remitting (RR) and Secondary Progressive are considered (SP). Two cohorts are considered: patients with a least 3 visits with EDSS in the last 3.25 years and patients with at least 6 visits with EDSS in the last 3.25 years.

### Appendix F.3. Performance per EDSS level at baseline

Tables F.10 and F.11 report the the ROC-AUC, AUC-PR, Brier Score and ECE by severity subgroup. Low severity patients are defined as the ones with *EDSS* ≤ 5.5 at baseline, while high severity patients are defined as having *EDSS >* 5.5 at baseline.

**Table F.7:** Summary statistics of the performance measures. Cohort with minimum 6 visits.

| Model | ROC-AUC | AUC-PR | Brier | ECE |
| --- | --- | --- | --- | --- |
| Ensemble | $0.71 \pm 0.02$ | $0.25 \pm 0.03$ | $0.11 \pm 0.01$ | $0.08 \pm 0.03$ |
| Attention | $0.7 \pm 0.02$ | $0.24 \pm 0.02$ | $0.11 \pm 0.01$ | $0.04 \pm 0.01$ |
| RNN | $0.7 \pm 0.02$ | $0.24 \pm 0.03$ | $0.11 \pm 0.01$ | $0.05 \pm 0.03$ |
| Static Bayesian NN | $0.68 \pm 0.02$ | $0.23 \pm 0.02$ | $0.11 \pm 0.01$ | $0.05 \pm 0.03$ |
| Dynamic Bayesian NN | $0.7 \pm 0.01$ | $0.24 \pm 0.02$ | $0.11 \pm 0.01$ | $0.08 \pm 0.02$ |
| Static Baseline | $0.67 \pm 0.02$ | $0.22 \pm 0.02$ | $0.11 \pm 0.01$ | $0.05 \pm 0.02$ |
| Dynamic Baseline | $0.69 \pm 0.02$ | $0.23 \pm 0.02$ | $0.11 \pm 0.01$ | $0.06 \pm 0.03$ |
| Static Logistic | $0.67 \pm 0.02$ | $0.22 \pm 0.02$ | $0.11 \pm 0.01$ | $0.04 \pm 0.02$ |
| Dynamic Logistic | $0.69 \pm 0.02$ | $0.25 \pm 0.02$ | $0.11 \pm 0.01$ | $0.04 \pm 0.03$ |
| Static DeepMTP | $0.66 \pm 0.03$ | $0.21 \pm 0.04$ | $0.12 \pm 0.02$ | $0.24 \pm 0.08$ |
| Dynamic DeepMTP | $0.67 \pm 0.03$ | $0.22 \pm 0.04$ | $0.12 \pm 0.01$ | $0.24 \pm 0.09$ |
| Static FactorizationMachine | $0.67 \pm 0.02$ | $0.22 \pm 0.02$ | $0.11 \pm 0.01$ | $0.13 \pm 0.02$ |
| Dynamic FactorizationMachine | $0.69 \pm 0.02$ | $0.24 \pm 0.03$ | $0.11 \pm 0.01$ | $0.15 \pm 0.05$ |

Two cohorts are considered: patients with a least 3 visits with EDSS in the last 3.25 years and patients with at least 6 visits with EDSS in the last 3.25 years.

## Appendix G. Machine Learning Models Details

### Appendix G.1. Bayesian Neural Networks

Introduced by Gal et al. [20], Monte Carlo Dropout is an approximate ensemble method for Bayesian Neural Networks (Bayesian NN or BNN). They prove that a Neural Network with dropout layers and L2-regularization approximates the predictive posterior distribution of a Gaussian process for a given data set. The resulting ensemble is in general well-calibrated [26], and more accurate than its non-Bayesian counterpart due to the regularizing effect. The Bayesian NN implemented in this work is identical to the baseline neural network previously introduced, with a few key differences. First, dropout [27] is applied between every layer. Second, the logits of the network are modeled as Gaussian distributions, rather than point estimates.

**Table F.8:**
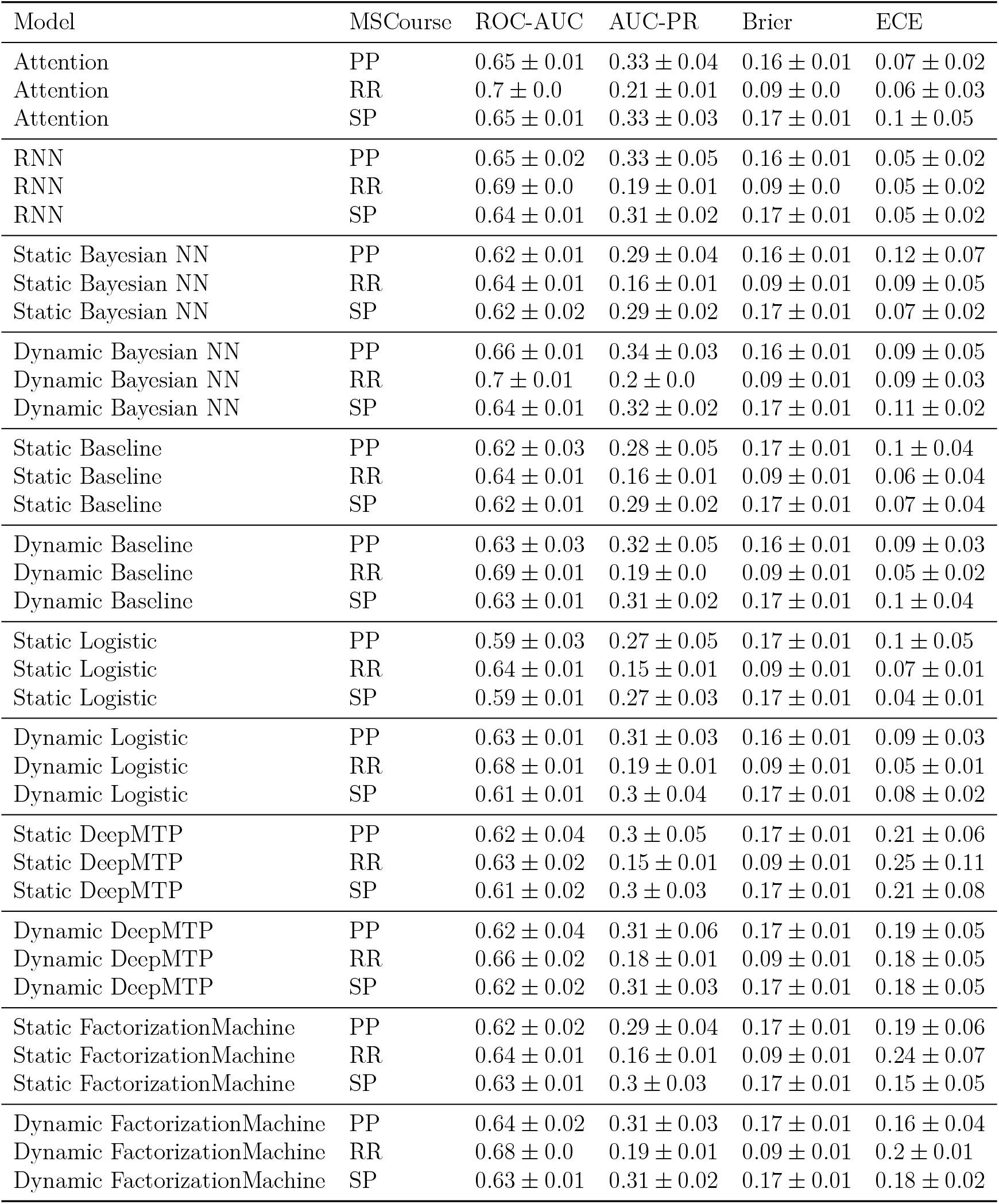
Results for disability progression prediction per MSCourse. Cohort with minimum 3 visits.

| Model | MSCourse | ROC-AUC | AUC-PR | Brier | ECE |
| --- | --- | --- | --- | --- | --- |
| Attention | PP | $0.65 \pm 0.01$ | $0.33 \pm 0.04$ | $0.16 \pm 0.01$ | $0.07 \pm 0.02$ |
| Attention | RR | $0.7 \pm 0.0$ | $0.21 \pm 0.01$ | $0.09 \pm 0.0$ | $0.06 \pm 0.03$ |
| Attention | SP | $0.65 \pm 0.01$ | $0.33 \pm 0.03$ | $0.17 \pm 0.01$ | $0.1 \pm 0.05$ |
| RNN | PP | $0.65 \pm 0.02$ | $0.33 \pm 0.05$ | $0.16 \pm 0.01$ | $0.05 \pm 0.02$ |
| RNN | RR | $0.69 \pm 0.0$ | $0.19 \pm 0.01$ | $0.09 \pm 0.0$ | $0.05 \pm 0.02$ |
| RNN | SP | $0.64 \pm 0.01$ | $0.31 \pm 0.02$ | $0.17 \pm 0.01$ | $0.05 \pm 0.02$ |
| Static Bayesian NN | PP | $0.62 \pm 0.01$ | $0.29 \pm 0.04$ | $0.16 \pm 0.01$ | $0.12 \pm 0.07$ |
| Static Bayesian NN | RR | $0.64 \pm 0.01$ | $0.16 \pm 0.01$ | $0.09 \pm 0.01$ | $0.09 \pm 0.05$ |
| Static Bayesian NN | SP | $0.62 \pm 0.02$ | $0.29 \pm 0.02$ | $0.17 \pm 0.01$ | $0.07 \pm 0.02$ |
| Dynamic Bayesian NN | PP | $0.66 \pm 0.01$ | $0.34 \pm 0.03$ | $0.16 \pm 0.01$ | $0.09 \pm 0.05$ |
| Dynamic Bayesian NN | RR | $0.7 \pm 0.01$ | $0.2 \pm 0.0$ | $0.09 \pm 0.01$ | $0.09 \pm 0.03$ |
| Dynamic Bayesian NN | SP | $0.64 \pm 0.01$ | $0.32 \pm 0.02$ | $0.17 \pm 0.01$ | $0.11 \pm 0.02$ |
| Static Baseline | PP | $0.62 \pm 0.03$ | $0.28 \pm 0.05$ | $0.17 \pm 0.01$ | $0.1 \pm 0.04$ |
| Static Baseline | RR | $0.64 \pm 0.01$ | $0.16 \pm 0.01$ | $0.09 \pm 0.01$ | $0.06 \pm 0.04$ |
| Static Baseline | SP | $0.62 \pm 0.01$ | $0.29 \pm 0.02$ | $0.17 \pm 0.01$ | $0.07 \pm 0.04$ |
| Dynamic Baseline | PP | $0.63 \pm 0.03$ | $0.32 \pm 0.05$ | $0.16 \pm 0.01$ | $0.09 \pm 0.03$ |
| Dynamic Baseline | RR | $0.69 \pm 0.01$ | $0.19 \pm 0.0$ | $0.09 \pm 0.01$ | $0.05 \pm 0.02$ |
| Dynamic Baseline | SP | $0.63 \pm 0.01$ | $0.31 \pm 0.02$ | $0.17 \pm 0.01$ | $0.1 \pm 0.04$ |
| Static Logistic | PP | $0.59 \pm 0.03$ | $0.27 \pm 0.05$ | $0.17 \pm 0.01$ | $0.1 \pm 0.05$ |
| Static Logistic | RR | $0.64 \pm 0.01$ | $0.15 \pm 0.01$ | $0.09 \pm 0.01$ | $0.07 \pm 0.01$ |
| Static Logistic | SP | $0.59 \pm 0.01$ | $0.27 \pm 0.03$ | $0.17 \pm 0.01$ | $0.04 \pm 0.01$ |
| Dynamic Logistic | PP | $0.63 \pm 0.01$ | $0.31 \pm 0.03$ | $0.16 \pm 0.01$ | $0.09 \pm 0.03$ |
| Dynamic Logistic | RR | $0.68 \pm 0.01$ | $0.19 \pm 0.01$ | $0.09 \pm 0.01$ | $0.05 \pm 0.01$ |
| Dynamic Logistic | SP | $0.61 \pm 0.01$ | $0.3 \pm 0.04$ | $0.17 \pm 0.01$ | $0.08 \pm 0.02$ |
| Static DeepMTP | PP | $0.62 \pm 0.04$ | $0.3 \pm 0.05$ | $0.17 \pm 0.01$ | $0.21 \pm 0.06$ |
| Static DeepMTP | RR | $0.63 \pm 0.02$ | $0.15 \pm 0.01$ | $0.09 \pm 0.01$ | $0.25 \pm 0.11$ |
| Static DeepMTP | SP | $0.61 \pm 0.02$ | $0.3 \pm 0.03$ | $0.17 \pm 0.01$ | $0.21 \pm 0.08$ |
| Dynamic DeepMTP | PP | $0.62 \pm 0.04$ | $0.31 \pm 0.06$ | $0.17 \pm 0.01$ | $0.19 \pm 0.05$ |
| Dynamic DeepMTP | RR | $0.66 \pm 0.02$ | $0.18 \pm 0.01$ | $0.09 \pm 0.01$ | $0.18 \pm 0.05$ |
| Dynamic DeepMTP | SP | $0.62 \pm 0.02$ | $0.31 \pm 0.03$ | $0.17 \pm 0.01$ | $0.18 \pm 0.05$ |
| Static FactorizationMachine | PP | $0.62 \pm 0.02$ | $0.29 \pm 0.04$ | $0.17 \pm 0.01$ | $0.19 \pm 0.06$ |
| Static FactorizationMachine | RR | $0.64 \pm 0.01$ | $0.16 \pm 0.01$ | $0.09 \pm 0.01$ | $0.24 \pm 0.07$ |
| Static FactorizationMachine | SP | $0.63 \pm 0.01$ | $0.3 \pm 0.03$ | $0.17 \pm 0.01$ | $0.15 \pm 0.05$ |
| Dynamic FactorizationMachine | PP | $0.64 \pm 0.02$ | $0.31 \pm 0.03$ | $0.17 \pm 0.01$ | $0.16 \pm 0.04$ |
| Dynamic FactorizationMachine | RR | $0.68 \pm 0.0$ | $0.19 \pm 0.01$ | $0.09 \pm 0.01$ | $0.2 \pm 0.01$ |
| Dynamic FactorizationMachine | SP | $0.63 \pm 0.01$ | $0.31 \pm 0.02$ | $0.17 \pm 0.01$ | $0.18 \pm 0.02$ |

**Table F.9:** Results for disability progression prediction per MSCourse. Cohort with minimum 6 visits

| Model | MSCourse | ROC-AUC | AUC-PR | Brier | ECE |
| --- | --- | --- | --- | --- | --- |
| Attention | PP | $0.62 \pm 0.03$ | $0.36 \pm 0.04$ | $0.19 \pm 0.01$ | $0.07 \pm 0.02$ |
| Attention | RR | $0.68 \pm 0.01$ | $0.18 \pm 0.02$ | $0.09 \pm 0.01$ | $0.07 \pm 0.03$ |
| Attention | SP | $0.61 \pm 0.02$ | $0.32 \pm 0.02$ | $0.18 \pm 0.02$ | $0.08 \pm 0.03$ |
| RNN | PP | $0.62 \pm 0.05$ | $0.36 \pm 0.05$ | $0.19 \pm 0.01$ | $0.09 \pm 0.03$ |
| RNN | RR | $0.68 \pm 0.02$ | $0.18 \pm 0.02$ | $0.09 \pm 0.01$ | $0.05 \pm 0.03$ |
| RNN | SP | $0.62 \pm 0.02$ | $0.32 \pm 0.03$ | $0.18 \pm 0.02$ | $0.07 \pm 0.01$ |
| Static Bayesian NN | PP | $0.63 \pm 0.05$ | $0.36 \pm 0.05$ | $0.19 \pm 0.01$ | $0.08 \pm 0.05$ |
| Static Bayesian NN | RR | $0.65 \pm 0.02$ | $0.16 \pm 0.01$ | $0.09 \pm 0.01$ | $0.08 \pm 0.03$ |
| Static Bayesian NN | SP | $0.59 \pm 0.05$ | $0.29 \pm 0.02$ | $0.18 \pm 0.02$ | $0.09 \pm 0.02$ |
| Dynamic Bayesian NN | PP | $0.63 \pm 0.04$ | $0.37 \pm 0.05$ | $0.19 \pm 0.01$ | $0.11 \pm 0.04$ |
| Dynamic Bayesian NN | RR | $0.68 \pm 0.01$ | $0.18 \pm 0.01$ | $0.09 \pm 0.01$ | $0.1 \pm 0.01$ |
| Dynamic Bayesian NN | SP | $0.61 \pm 0.04$ | $0.31 \pm 0.02$ | $0.18 \pm 0.02$ | $0.12 \pm 0.03$ |
| Static Baseline | PP | $0.62 \pm 0.04$ | $0.35 \pm 0.04$ | $0.19 \pm 0.01$ | $0.09 \pm 0.03$ |
| Static Baseline | RR | $0.64 \pm 0.02$ | $0.15 \pm 0.01$ | $0.09 \pm 0.01$ | $0.05 \pm 0.03$ |
| Static Baseline | SP | $0.59 \pm 0.03$ | $0.29 \pm 0.02$ | $0.18 \pm 0.02$ | $0.07 \pm 0.01$ |
| Dynamic Baseline | PP | $0.62 \pm 0.05$ | $0.35 \pm 0.07$ | $0.19 \pm 0.01$ | $0.09 \pm 0.03$ |
| Dynamic Baseline | RR | $0.67 \pm 0.02$ | $0.17 \pm 0.01$ | $0.09 \pm 0.01$ | $0.08 \pm 0.03$ |
| Dynamic Baseline | SP | $0.61 \pm 0.03$ | $0.32 \pm 0.02$ | $0.18 \pm 0.02$ | $0.08 \pm 0.02$ |
| Static Logistic | PP | $0.61 \pm 0.03$ | $0.35 \pm 0.02$ | $0.19 \pm 0.01$ | $0.1 \pm 0.05$ |
| Static Logistic | RR | $0.63 \pm 0.02$ | $0.15 \pm 0.01$ | $0.09 \pm 0.01$ | $0.07 \pm 0.03$ |
| Static Logistic | SP | $0.58 \pm 0.03$ | $0.29 \pm 0.02$ | $0.18 \pm 0.02$ | $0.07 \pm 0.03$ |
| Dynamic Logistic | PP | $0.64 \pm 0.03$ | $0.38 \pm 0.02$ | $0.18 \pm 0.01$ | $0.12 \pm 0.05$ |
| Dynamic Logistic | RR | $0.67 \pm 0.01$ | $0.19 \pm 0.01$ | $0.09 \pm 0.01$ | $0.06 \pm 0.02$ |
| Dynamic Logistic | SP | $0.6 \pm 0.03$ | $0.31 \pm 0.02$ | $0.18 \pm 0.02$ | $0.07 \pm 0.04$ |
| Static DeepMTP | PP | $0.61 \pm 0.06$ | $0.36 \pm 0.04$ | $0.19 \pm 0.02$ | $0.19 \pm 0.04$ |
| Static DeepMTP | RR | $0.63 \pm 0.02$ | $0.15 \pm 0.02$ | $0.1 \pm 0.02$ | $0.22 \pm 0.11$ |
| Static DeepMTP | SP | $0.6 \pm 0.03$ | $0.29 \pm 0.04$ | $0.19 \pm 0.01$ | $0.22 \pm 0.04$ |
| Dynamic DeepMTP | PP | $0.6 \pm 0.05$ | $0.34 \pm 0.03$ | $0.2 \pm 0.02$ | $0.21 \pm 0.07$ |
| Dynamic DeepMTP | RR | $0.65 \pm 0.03$ | $0.17 \pm 0.04$ | $0.1 \pm 0.02$ | $0.24 \pm 0.1$ |
| Dynamic DeepMTP | SP | $0.6 \pm 0.02$ | $0.31 \pm 0.03$ | $0.19 \pm 0.01$ | $0.22 \pm 0.07$ |
| Static FactorizationMachine | PP | $0.62 \pm 0.03$ | $0.36 \pm 0.04$ | $0.19 \pm 0.02$ | $0.14 \pm 0.03$ |
| Static FactorizationMachine | RR | $0.63 \pm 0.02$ | $0.15 \pm 0.02$ | $0.09 \pm 0.01$ | $0.16 \pm 0.07$ |
| Static FactorizationMachine | SP | $0.6 \pm 0.03$ | $0.31 \pm 0.03$ | $0.18 \pm 0.02$ | $0.14 \pm 0.04$ |
| Dynamic FactorizationMachine | PP | $0.63 \pm 0.02$ | $0.37 \pm 0.05$ | $0.19 \pm 0.01$ | $0.16 \pm 0.06$ |
| Dynamic FactorizationMachine | RR | $0.66 \pm 0.01$ | $0.18 \pm 0.02$ | $0.09 \pm 0.01$ | $0.15 \pm 0.05$ |
| Dynamic FactorizationMachine | SP | $0.61 \pm 0.01$ | $0.31 \pm 0.03$ | $0.19 \pm 0.02$ | $0.18 \pm 0.04$ |

**Table F.10:** Results for disability progression prediction for EDSS ≤ 5.5 and *>* 5.5. Cohort with minimum 3 visits

| Model | EDSS | ROC-AUC | AUC-PR | Brier | ECE |
| --- | --- | --- | --- | --- | --- |
| Attention | $EDSS \leq 5.5$ | $0.72 \pm 0.01$ | $0.26 \pm 0.01$ | $0.09 \pm 0.0$ | $0.07 \pm 0.04$ |
| Attention | $EDSS > 5.5$ | $0.65 \pm 0.01$ | $0.27 \pm 0.04$ | $0.15 \pm 0.01$ | $0.07 \pm 0.02$ |
| RNN | $EDSS \leq 5.5$ | $0.71 \pm 0.01$ | $0.24 \pm 0.02$ | $0.1 \pm 0.0$ | $0.04 \pm 0.03$ |
| RNN | $EDSS > 5.5$ | $0.64 \pm 0.01$ | $0.26 \pm 0.03$ | $0.15 \pm 0.02$ | $0.05 \pm 0.03$ |
| Static Bayesian NN | $EDSS \leq 5.5$ | $0.67 \pm 0.01$ | $0.21 \pm 0.01$ | $0.1 \pm 0.0$ | $0.1 \pm 0.04$ |
| Static Bayesian NN | $EDSS > 5.5$ | $0.63 \pm 0.01$ | $0.25 \pm 0.03$ | $0.15 \pm 0.02$ | $0.09 \pm 0.04$ |
| Dynamic Bayesian NN | $EDSS \leq 5.5$ | $0.72 \pm 0.01$ | $0.25 \pm 0.01$ | $0.09 \pm 0.0$ | $0.08 \pm 0.04$ |
| Dynamic Bayesian NN | $EDSS > 5.5$ | $0.64 \pm 0.02$ | $0.26 \pm 0.03$ | $0.15 \pm 0.01$ | $0.11 \pm 0.03$ |
| Static Baseline | $EDSS \leq 5.5$ | $0.67 \pm 0.01$ | $0.2 \pm 0.02$ | $0.1 \pm 0.01$ | $0.07 \pm 0.05$ |
| Static Baseline | $EDSS > 5.5$ | $0.63 \pm 0.01$ | $0.25 \pm 0.03$ | $0.15 \pm 0.02$ | $0.09 \pm 0.04$ |
| Dynamic Baseline | $EDSS \leq 5.5$ | $0.71 \pm 0.01$ | $0.24 \pm 0.01$ | $0.1 \pm 0.01$ | $0.09 \pm 0.03$ |
| Dynamic Baseline | $EDSS > 5.5$ | $0.63 \pm 0.01$ | $0.26 \pm 0.04$ | $0.15 \pm 0.02$ | $0.09 \pm 0.03$ |
| Static Logistic | $EDSS \leq 5.5$ | $0.66 \pm 0.01$ | $0.2 \pm 0.01$ | $0.1 \pm 0.01$ | $0.03 \pm 0.01$ |
| Static Logistic | $EDSS > 5.5$ | $0.61 \pm 0.02$ | $0.24 \pm 0.04$ | $0.15 \pm 0.01$ | $0.05 \pm 0.01$ |
| Dynamic Logistic | $EDSS \leq 5.5$ | $0.7 \pm 0.0$ | $0.24 \pm 0.02$ | $0.1 \pm 0.0$ | $0.06 \pm 0.04$ |
| Dynamic Logistic | $EDSS > 5.5$ | $0.62 \pm 0.01$ | $0.25 \pm 0.04$ | $0.15 \pm 0.01$ | $0.07 \pm 0.03$ |
| Static DeepMTP | $EDSS \leq 5.5$ | $0.65 \pm 0.02$ | $0.19 \pm 0.02$ | $0.1 \pm 0.0$ | $0.23 \pm 0.06$ |
| Static DeepMTP | $EDSS > 5.5$ | $0.63 \pm 0.02$ | $0.25 \pm 0.04$ | $0.15 \pm 0.02$ | $0.21 \pm 0.05$ |
| Dynamic DeepMTP | $EDSS \leq 5.5$ | $0.69 \pm 0.02$ | $0.22 \pm 0.02$ | $0.1 \pm 0.0$ | $0.18 \pm 0.03$ |
| Dynamic DeepMTP | $EDSS > 5.5$ | $0.62 \pm 0.02$ | $0.26 \pm 0.04$ | $0.15 \pm 0.02$ | $0.19 \pm 0.08$ |
| Static FactorizationMachine | $EDSS \leq 5.5$ | $0.67 \pm 0.01$ | $0.21 \pm 0.02$ | $0.1 \pm 0.0$ | $0.18 \pm 0.06$ |
| Static FactorizationMachine | $EDSS > 5.5$ | $0.63 \pm 0.01$ | $0.26 \pm 0.04$ | $0.15 \pm 0.02$ | $0.18 \pm 0.06$ |
| Dynamic FactorizationMachine | $EDSS \leq 5.5$ | $0.7 \pm 0.01$ | $0.23 \pm 0.01$ | $0.1 \pm 0.0$ | $0.14 \pm 0.03$ |
| Dynamic FactorizationMachine | $EDSS > 5.5$ | $0.64 \pm 0.01$ | $0.26 \pm 0.03$ | $0.15 \pm 0.02$ | $0.22 \pm 0.05$ |

**Table F.11:** Results for disability progression prediction for EDSS ≤ 5.5 and *>* 5.5. Cohort with minimum 6 visits

| Model | EDSS | ROC-AUC | AUC-PR | Brier | ECE |
| --- | --- | --- | --- | --- | --- |
| Attention | $EDSS \leq 5.5$ | $0.7 \pm 0.02$ | $0.23 \pm 0.03$ | $0.09 \pm 0.01$ | $0.05 \pm 0.03$ |
| Attention | $EDSS > 5.5$ | $0.62 \pm 0.01$ | $0.29 \pm 0.01$ | $0.17 \pm 0.01$ | $0.07 \pm 0.04$ |
| RNN | $EDSS \leq 5.5$ | $0.7 \pm 0.02$ | $0.22 \pm 0.03$ | $0.09 \pm 0.01$ | $0.08 \pm 0.03$ |
| RNN | $EDSS > 5.5$ | $0.62 \pm 0.01$ | $0.28 \pm 0.01$ | $0.17 \pm 0.01$ | $0.06 \pm 0.03$ |
| Static Bayesian NN | $EDSS \leq 5.5$ | $0.67 \pm 0.02$ | $0.2 \pm 0.02$ | $0.1 \pm 0.01$ | $0.07 \pm 0.05$ |
| Static Bayesian NN | $EDSS > 5.5$ | $0.62 \pm 0.03$ | $0.28 \pm 0.02$ | $0.17 \pm 0.01$ | $0.06 \pm 0.02$ |
| Dynamic Bayesian NN | $EDSS \leq 5.5$ | $0.7 \pm 0.01$ | $0.23 \pm 0.02$ | $0.09 \pm 0.01$ | $0.1 \pm 0.02$ |
| Dynamic Bayesian NN | $EDSS > 5.5$ | $0.61 \pm 0.02$ | $0.28 \pm 0.01$ | $0.17 \pm 0.01$ | $0.08 \pm 0.04$ |
| Static Baseline | $EDSS \leq 5.5$ | $0.66 \pm 0.02$ | $0.19 \pm 0.02$ | $0.1 \pm 0.01$ | $0.04 \pm 0.02$ |
| Static Baseline | $EDSS > 5.5$ | $0.61 \pm 0.03$ | $0.28 \pm 0.01$ | $0.17 \pm 0.01$ | $0.07 \pm 0.03$ |
| Dynamic Baseline | $EDSS \leq 5.5$ | $0.69 \pm 0.02$ | $0.22 \pm 0.03$ | $0.09 \pm 0.01$ | $0.07 \pm 0.03$ |
| Dynamic Baseline | $EDSS > 5.5$ | $0.61 \pm 0.02$ | $0.28 \pm 0.02$ | $0.17 \pm 0.01$ | $0.07 \pm 0.02$ |
| Static Logistic | $EDSS \leq 5.5$ | $0.66 \pm 0.02$ | $0.19 \pm 0.02$ | $0.1 \pm 0.01$ | $0.05 \pm 0.03$ |
| Static Logistic | $EDSS > 5.5$ | $0.6 \pm 0.02$ | $0.27 \pm 0.02$ | $0.17 \pm 0.01$ | $0.05 \pm 0.01$ |
| Dynamic Logistic | $EDSS \leq 5.5$ | $0.69 \pm 0.02$ | $0.23 \pm 0.02$ | $0.09 \pm 0.01$ | $0.05 \pm 0.03$ |
| Dynamic Logistic | $EDSS > 5.5$ | $0.61 \pm 0.01$ | $0.28 \pm 0.01$ | $0.17 \pm 0.01$ | $0.06 \pm 0.05$ |
| Static DeepMTP | $EDSS \leq 5.5$ | $0.65 \pm 0.03$ | $0.19 \pm 0.04$ | $0.11 \pm 0.02$ | $0.24 \pm 0.1$ |
| Static DeepMTP | $EDSS > 5.5$ | $0.62 \pm 0.02$ | $0.28 \pm 0.02$ | $0.17 \pm 0.0$ | $0.19 \pm 0.06$ |
| Dynamic DeepMTP | $EDSS \leq 5.5$ | $0.67 \pm 0.02$ | $0.2 \pm 0.04$ | $0.11 \pm 0.02$ | $0.23 \pm 0.1$ |
| Dynamic DeepMTP | $EDSS > 5.5$ | $0.62 \pm 0.02$ | $0.28 \pm 0.01$ | $0.17 \pm 0.0$ | $0.26 \pm 0.05$ |
| Static FactorizationMachine | $EDSS \leq 5.5$ | $0.66 \pm 0.02$ | $0.2 \pm 0.02$ | $0.1 \pm 0.01$ | $0.13 \pm 0.02$ |
| Static FactorizationMachine | $EDSS > 5.5$ | $0.61 \pm 0.03$ | $0.28 \pm 0.02$ | $0.17 \pm 0.01$ | $0.13 \pm 0.04$ |
| Dynamic FactorizationMachine | $EDSS \leq 5.5$ | $0.69 \pm 0.02$ | $0.22 \pm 0.03$ | $0.09 \pm 0.01$ | $0.11 \pm 0.04$ |
| Dynamic FactorizationMachine | $EDSS > 5.5$ | $0.6 \pm 0.01$ | $0.27 \pm 0.01$ | $0.17 \pm 0.01$ | $0.2 \pm 0.05$ |

Third, the loss function is a modified cross-entropy loss, that samples from the aforementioned logit distributions [26]. While Kendall et al. [26] define a loss that allows capturing aleatoric uncertainty, we simplify it further to work for our binary classification. We use Monte Carlo integration to approximate the distribution.

The resulting network has a way of expressing epistemic (model-bound) uncertainty by using dropout, creating a distribution over the model weights. On the other hand, the resulting BNN also has a way of expressing aleatoric uncertainty, by modelling the output of the network as a Gaussian distribution in logit space.

We define the network to have two outputs, *µ* and log *σ*^2^. Instead of learning variance, log variance is learnt to constrain the value to be positive. Our loss function for a single input, with *T* the amount of Monte Carlo integration samples, and *y* the true label is defined as follows:

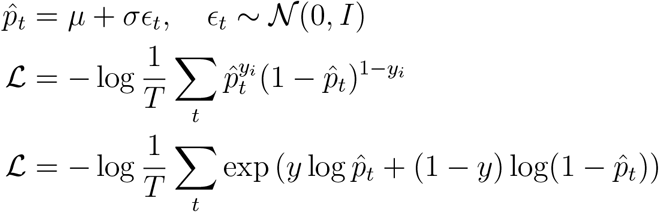

### Appendix G.2. DeepMTP

The DeepMTP framework was introduced by Iliadis et al. [28] as a unified approach for multi-target prediction (MTP) problems. Applications that fall under the umbrella of MTP are concerned with the simultaneous prediction of multiple target variables. Even though this work focuses on the prediction of a single binary variable (patient progresses or not), we are able to use the DeepMTP framework by applying a multi-task trick. To achieve this, we select one categorical feature from the available data set (country) and create multiple targets (or tasks), thus forming a multi-task learning problem. By doing this, the goal becomes the prediction of the progression of a patient depending on the country (s)he is residing in. Even though the natural benchmark comparison of a multi-task problem is a collection of models that are trained on subsets of the original data set belonging to a single country (single-task models), we believe that this is out of the scope of this work. For this reason, after prediction, we collapse the tasks and create a single prediction that is comparable with the other methods tested in this paper.

In terms of architecture, DeepMTP uses a two-branch architecture that is flexible enough to be adapted for the different MTP prediction settings. In this specific multi-task case, the first branch encodes the same features as all the other methods, and the second uses a one-hot encoded vector that maps to a given country. Both branches are comprised of one or more fully connected layers, and their outputs are combined using a dot product. The single value resulting from the dot product is the output of the entire network (progression or not).

### Appendix G.3. Factorization Machines

In 2010, Rendle introduced Factorization Machines (FM) as a new model class that combines the advantages of Support Vector Machines (SVM) with factorization models [29]. Factorization machines model interactions between features using factorized parameters. The prediction function of degree two, meaning that pairwise interactions represent the highest degree of interaction considered, is given by:

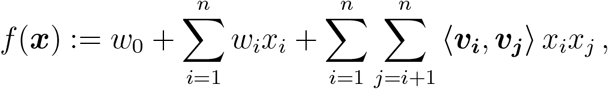

with model parameters *w*_0_ ∈ ℝ, ***w*** ∈ ℝ^*n*^ and ***V*** ∈ ℝ^*n×k*^, and the dot product 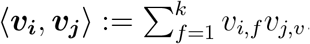.

Factorization Machines are more widely applicable than regular factorization models such as matrix factorization, as they naturally include features as well and learn to map them into a lower-dimensional latent factor space. This behaviour explains why FM can be surprisingly successful when working with categorical features (e.g. country, sex), even under high sparsity. Also, thanks to their linear time complexity, they are often applied in large real-world recommendation data sets [30].

In this work, we used stochastic gradient descent with adaptive regularization as a learning method [31, 30]. A Python implementation is available at https://github.com/godpgf/pylibfm. The main hyper-parameter to tune is the size *k* of the latent factor space.

## Appendix H. Supplementary information on variables used as predictors

### Appendix H.1. Information on treatments

Except for Mitoxantrone and Cyclophosphamide, we assumed that only one DMT was administered at the same time. This implies that if a new DMT was started, the administration of the previous DMT was considered to have ended, even if no end date was registered in the data. Mitoxantrone and Cyclophosphamide can be administered in combination with another DMT. Indeed, they are induction DMTs and are thus expected to have a long-term effect. Therefore, only the start dates of these two DMTs were recorded. They were coded by a separate category: highly active induction DMTs. Alemtuzumab and Cladribine are also induction DMTs. In contrast to Mitoxantrone and Cyclophosphamide they are not combined with other DMTs. If a new DMT was started, it was assumed that they were considered as not effective and the start date of the new DMT was taken as the end date of Alemtuzumab or Cladribine.

### Appendix H.2. Grouping of the included clinical variables

The *static feature set* contains variables available at *t* = 0 without taking into account possible previous values. Categorical variables can be encoded as indicator variables. For example, sex is encoded as female yes / no and male yes / no. If data can be missing, the category ”unknown” is added. EDSS and the KFS scores were treated as continuous variables, even though they are categorical. The variables of the static feature set are: Sex, Age (years), Age at MS onset (years), Disease duration (years), MS course at *t* = 0 (RRMS, SPMS, PPMS), EDSS at *t* = 0, Last used DMT at *t* = 0, Use of induction DMT at *t* = 0, all KFS scores at *t* = 0, education status, first symptom (supratentorial, optic pathways, brainstem, spinal cord or missing), time of prediction (years since 1990), time of diagnosis (years since 1990).

The *dynamic feature set* adds information about behaviour before *t* = 0 (longitudinal information) to the static data set. It contains variables that are hand-engineered from the longitudinal variables: number of visits in the last 3.25 years, the minimum and maximum in the whole history (*t* ≤ 0) of the EDSS and all KFS variables, mean and standard deviation over the last 3.25 years of the EDSS and all KFS scores, oldest EDSS and KFS score measured in the last 3.25 years, relapse rate over the whole history (number of relapses divided by the follow-up period - since first clinical visit), time since the last relapse (years), presence of high-efficacy DMT in the past, disease duration until a first DMT was administered, disease duration until an high-efficacy active DMT was administrated, time spent on a DMT during the disease duration (ratio of time on a DMT divided by the time since MS onset), time since the last Fampridine administration

The variables time since the last relapse, disease duration until a DMT was administered, disease duration until a high-efficacy DMT was administrated and time since the last Fampridine administration are transformed according to an 1*/*(1 + *t*) scaling, with *t* the actual time. If no time can be defined because, e.g., no DMT has ever been administered, the transformed variable is 0. If *t <* 0, which can happen because of erroneous dates in the data set, the transformed variable is set to 1.

The *longitudinal feature set* contains the dates and values for the following variables: all measured EDSS values and KFS scores, relapses occurrence (encoded as a binary variable set to 1 when a relapse occurs), relapse position (brainstem, pyramidal tract or other), cumulative relapse count, MS course, DMT administration (start and end dates), induction DMT administration (start date), Fampridine administration. The timing of measurements is expected to be informative [25, 17, 32].

## Appendix I. Extra information on target definition

Importantly, if progression *w* = 1 cannot be confirmed because there are no EDSS measurements after two years that can be used for confirmation, it is not a valid target and no label can be derived. If progression cannot be confirmed because an EDSS used for confirmation leads to *w* = 0, it counts as no disability progression (*w*_*c*_ = 0). If there is no disability progression (*w* = 0), no confirmation is needed to make it a valid target. Note that even with confirmation for at least six months, around 20% of progression events are expected to regress after more than five years [33]. However, disability progression that lasts several years is a relevant outcome for the person with MS.

Episodes were considered valid if they meet the following criteria. The time at which the prediction were made should be after 1990 (*t*_0_ *>* 1990, Jan 1st). This ensured that we had a cohort of patients from decades were disease modifying therapies (DMTs) were available [34].

All variables measured at visits after 1970 were used to perform the prediction. We further required a minimum number of EDSS measurements in the last three years and three months of the observation window. In this study, two cohorts are investigated, one with a minimum of three EDSS measurements, the other with a minimum of six EDSS measurements. This excluded patients who have a less than yearly (or biyearly) EDSS follow-up frequency. The three additional months were chosen to allow for some margin in when the yearly visit was planned. The patient should not be classified as being in a clinically isolated syndrome (CIS) at *t* = 0.

To summarize our target of disability progression in words: the patient will experience a disability progression event somewhere in the next two years. This event is sustained for at least six months and at least until two years after the time the prediction is made.

## Appendix J. List of MSBase authors

- Dana Horakova, Charles University in Prague and General University Hospital, Prague, Czech Republic
- Francesco Patti, Department of Medical and Surgical Sciences and Advanced Technologies, GF Ingrassia, Catania, Italy
- Guillermo Izquierdo, Hospital Universitario Virgen Macarena, Sevilla, Spain
- Sara Eichau, Hospital Universitario Virgen Macarena, Sevilla, Spain
- Alexandre Prat, CHUM and Université de Montreal, Montreal, Canada
- Alessandra Lugaresi, IRCCS Istituto delle Scienze Neurologiche di Bologna, Bologna, Italia and Dipartimento di Scienze Biomediche e Neuromotorie, Università di Bologna, Bologna, Italia
- Pierre Grammond, CISSS Chaudière-Appalache, Levis, Canada
- Tomas Kalincik, Melbourne MS Centre, Department of Neurology, Royal Melbourne Hospital, Melbourne, Australia
- Francois Grand’Maison, Neuro Rive-Sud, Quebec, Canada
- Olga Skibina, Box Hill Hospital, Melbourne, Australia
- Murat Terzi, 19 Mayis University, Samsun, Turkey

**Figure I.9:**
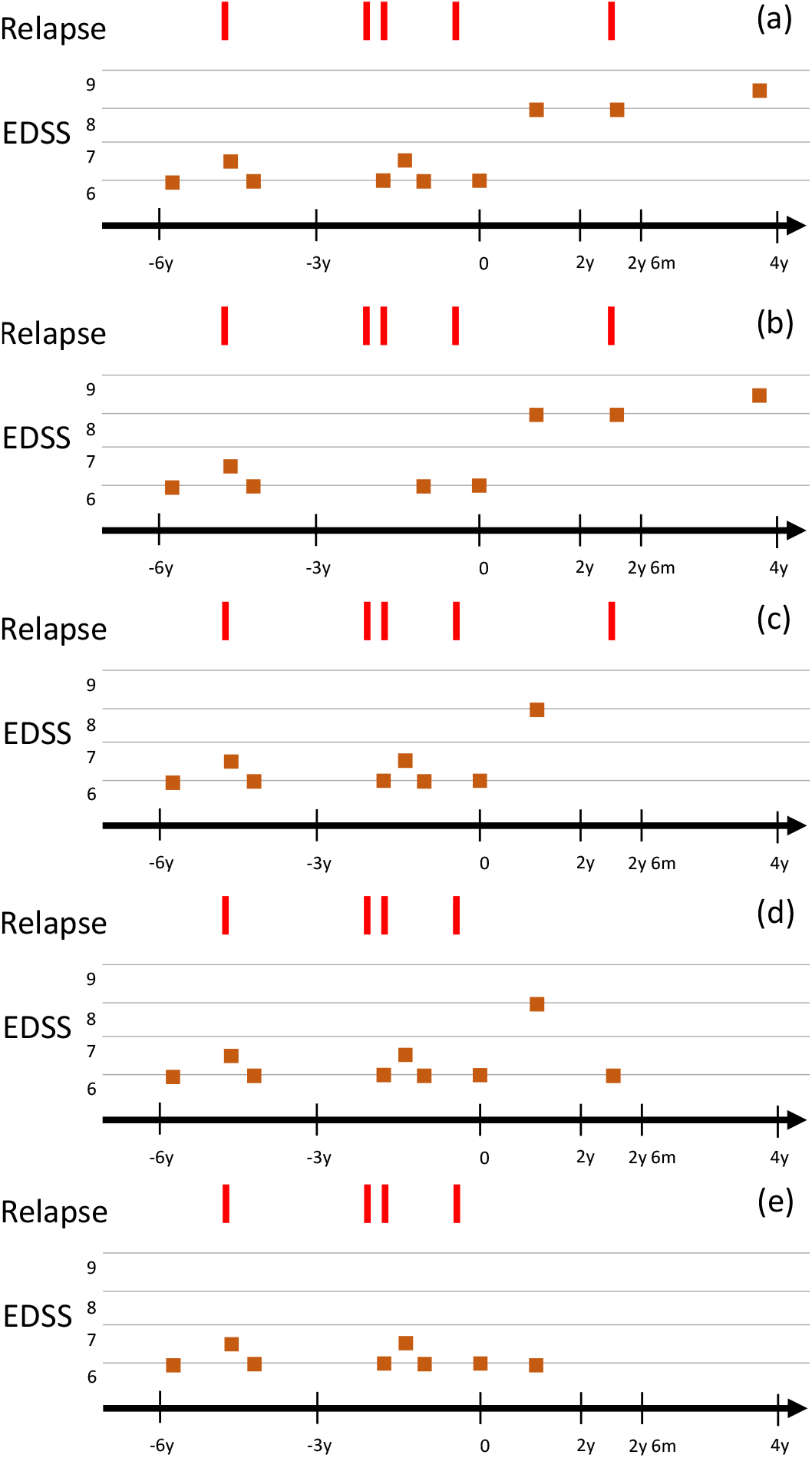
Examples of valid and non-valid samples. The time is in years (*y*) and months (*m*). (a) Confirmed progression after 2 years. The EDSS around 2*y*6*m* is not used to confirm the progression, because it occurs within 1 month after a relapse. Progression is confirmed with the EDSS measurement around 4*y*. There are 3 EDSS measurements between −3*y* and 0*y*, which is enough follow-up data. (b) This is not a valid sample: there are not enough EDSS measurements between −3*y* and 0*y*. (c) This is not a valid sample: no confirmed progression because there are4n0o EDSS values after 2*y*. (d) This is a valid sample: the EDSS decreases after 2*y*, so this counts as no disability progression. (e) This is a valid sample: *w*_*u*_ = 0, so no confirmation is needed.
- Maria Edite Rio, Centro Hospitalar Universitario de Sao Joao, Porto, Portugal
- Pamela McCombe, University of Queensland, Brisbane, Australia
- Mark Slee, Flinders University, Adelaide, Australia
- Saloua Mrabet, Razi Hospital, Manouba, Tunisia
- Jeannette Lechner-Scott, University Newcastle, Newcastle, Australia
- Oliver Gerlach, Academic MS Center Zuyderland, Department of Neurology, Zuyderland Medical Center, Sittard-Geleen, The Netherlands, and School for Mental Health and Neuroscience, Maastricht University, Maastricht, The Netherlands.
- Samia J. Khoury, American University of Beirut Medical Center, Beirut, Lebanon
- Elisabetta Cartechini, Azienda Sanitaria Unica Regionale Marche - AV3, Macerata, Italy
- Maria Jose Sa, Centro Hospitalar Universitario de Sao Joao, Porto, Portugal
- Vincent Van Pesch, Cliniques Universitaires Saint-Luc, Brussels, Belgium
- Bianca Weinstock-Guttman, Buffalo General Medical Center
- Yolanda Blanco, Hospital Clinic de Barcelona, Barcelona, Spain
- Radek Ampapa, Nemocnice Jihlava, Jihlava, Czech Republic
- Daniele Spitaleri, Azienda Ospedaliera di Rilievo Nazionale San Giuseppe Moscati Avellino, Avellino, Italy
- Claudio Solaro, Dept. of Rehabilitation, CRFF Mons. Luigi Novarese, Moncrivello, Italy
- Davide Maimone, MS center, UOC Neurologia, ARNAS Garibaldi, Catania, Italy.
- Aysun Soysal, Bakirkoy Education and Research Hospital for Psychiatric and Neurological Diseases, Istanbul, Turkey
- Gerardo Iuliano (retired - no PI successor but has approved ongoing use of data), Ospedali Riuniti di Salerno, Salerno, Italy
- Bart Van Wijmeersch, Rehabilitation and MS-Centre Overpelt and Hasselt University, Hasselt, Belgium
- Riadh Gouider, Razi Hospital, Manouba, Tunisia
- Tamara Castillo-Triviño, Hospital Universitario Donostia, San Sebastián, Spain
- Jose Luis Sanchez-Menoyo, Hospital de Galdakao-Usansolo, Galdakao, Spain
- Guy Laureys, Universitary Hospital Ghent, Ghent, Belgium
- Anneke van der Walt, The Alfred Hospital, Melbourne, Australia
- Jiwon Oh, St. Michael’s Hospital, Toronto, Canada
- Eduardo Aguera-Morales, University Hospital Reina Sofia, Cordoba, Spain
- Ayse Altintas, Koc University, School of Medicine, Istanbul, Turkey
- Abdullah Al-Asmi, College of Medicine & Health Sciences and Sultan Qaboos University Hospital, SQU, Oman
- Koen de Gans, Groene Hart Ziekenhuis, Gouda, Netherlands
- Yara Fragoso, Universidade Metropolitana de Santos, Santos, Brazil
- Tunde Csepany, University of Debrecen, Debrecen, Hungary
- Suzanne Hodgkinson, Liverpool Hospital, Sydney, Australia
- Norma Deri, Hospital Fernandez, Capital Federal, Argentina
- Talal Al-Harbi, King Fahad Specialist Hospital-Dammam, Khobar, Saudi Arabia
- Bruce Taylor, Royal Hobart Hospital, Hobart, Australia
- Orla Gray, South Eastern HSC Trust, Belfast, United Kingdom
- Patrice Lalive, Geneva University Hospital, Geneva, Switzerland
- Csilla Rozsa, Jahn Ferenc Teaching Hospital, Budapest, Hungary
- Chris McGuigan, St Vincent’s University Hospital, Dublin, Ireland
- Allan Kermode, University of Western Australia, Nedlands, Australia
- Angel Perez sempere, Hospital General Universitario de Alicante, Alicante, Spain
- Simu Mihaela, Emergency Clinical County Hospital ”Pius Brinzeu”, Timisoara, Romania and University of Medicine and Pharmacy Victor Babes, Timisoara, Romania.
- Magdolna Simo, Semmelweis University Budapest, Budapest, Hungary
- Todd Hardy, Concord Repatriation General Hospital, Sydney, Australia
- Danny Decoo, AZ Alma Ziekenhuis, Sijsele - Damme, Belgium
- Stella Hughes, Royal Victoria Hospital, Belfast, United Kingdom
- Norbert Vella, Mater Dei Hospital, Msida, Malta
- Attila Sas, BAZ County Hospital, Miskolc, Hungary
- Nikolaos Grigoriadis, AHEPA University Hospital, Thessaloniki, Greece

## Appendix K. List of MSBase contributors

- Eva Kubala Havrdova, Charles University in Prague and General University Hospital, Prague, Czech Republic
- Serkan Ozakbas, Dokuz Eylul University, Konak/Izmir, Turkey
- Marc Girard, CHUM and Université de Montreal, Montreal, Canada
- Marco Onofrj, University G. d’Annunzio, Chieti, Italy
- Raed Alroughani, Amiri Hospital, Sharq, Kuwait
- Maria Pia Amato, University of Florence, Florence, Italy
- Katherine Buzzard, Box Hill Hospital, Melbourne, Australia
- Cavit Boz, KTU Medical Faculty Farabi Hospital, Trabzon, Turkey
- Vahid Shaygannejad, Isfahan University of Medical Sciences, Isfahan, Iran
- Jens Kuhle, Universitatsspital Basel, Basel, Switzerland
- Bassem Yamout, American University of Beirut Medical Center, Beirut, Lebanon
- Recai Turkoglu, Haydarpasa Numune Training and Research Hospital, Istanbul, Turkey
- Julie Prevost, CSSS Saint-Jérôme, Saint-Jerome, Canada
- Ernest Butler, Monash Medical Centre, Melbourne, Australia
- Celia Oreja-Guevara, Hospital Clinico San Carlos, Madrid, Spain
- Richard Macdonell, Austin Health, Melbourne, Australia
- Ricardo Fernandez Bolaños, Hospital Universitario Virgen de Valme, Seville, Spain
- Marie D’hooghe, Nationaal MS Centrum, Melsbroek, Belgium
- Liesbeth Van Hijfte, Universitary Hospital Ghent, Ghent, Belgium
- Helmut Butzkueven, The Alfred Hospital, Melbourne, Australia
- Michael Barnett, Brain and Mind Centre, Sydney, Australia
- Justin Garber, Westmead Hospital, Sydney, Australia
- Sarah Besora, Hospital Universitari MútuaTerrassa, Barcelona, Spain
- Edgardo Cristiano, Centro de Esclerosis Múltiple de Buenos Aires (CEMBA), Buenos Aires, Argentina
- Magd Zakaria, Ain Shams University
- Maria Laura Saladino, INEBA - Institute of Neuroscience Buenos Aires, Buenos Aires, Argentina
- Shlomo Flechter, Assaf Harofeh Medical Center, Beer-Yaakov, Israel
- Leontien Den braber-Moerland, Francicus Ziekenhuis, Roosendaal, Netherlands
- Fraser Moore, Jewish General Hospital, Montreal, Canada
- Rana Karabudak, Hacettepe University, Ankara, Turkey
- Claudio Gobbi, Ospedale Civico Lugano, Lugano, Switzerland
- Jennifer Massey, St Vincent’s Hospital, Sydney, Australia
- Nevin Shalaby, Kasr Al Ainy MS research Unit (KAMSU), Cairo, Egypt
- Jabir Alkhaboori, Royal Hospital, Muscat, Oman
- Cameron Shaw, Geelong Hospital, Geelong, Australia
- Jose Andres Dominguez, Hospital Universitario de la Ribera, Alzira, Spain
- Jan Schepel, Waikato Hospital, Hamilton, New Zealand
- Krisztina Kovacs, Péterfy Sandor Hospital, Budapest, Hungary
- Pamela McCombe, Royal Brisbane and Women’s Hospital, Brisbane, Australia
- Bhim Singhal, Bombay Hospital Institute of Medical Sciences, Mumbai, India
- Mike Boggild, Townsville Hospital, Townsville, Australia
- Imre Piroska, Veszprém Megyei Csolnoky Ferenc Kórház zrt., Veszprem, Hungary
- Neil Shuey, St Vincents Hospital, Fitzroy, Melbourne, Australia
- Carlos Vrech, Sanatorio Allende, Cordoba, Argentina
- Tatjana Petkovska-Boskova, Clinic of Neurology Clinical Center, Skopje, Macedonia
- Ilya Kister, New York University Langone Medical Center, New York, United States
- Cees Zwanikken, University Hospital Nijmegen, Nijmegen, Netherlands
- Jamie Campbell, Craigavon Area Hospital, Craigavon, United Kingdom
- Etienne Roullet, MS Clinic, Hopital Tenon, Paris, France
- Cristina Ramo-Tello, Hospital Germans Trias i Pujol, Badalona, Spain
- Jose Antonio Cabrera-Gomez, Centro Internacional de Restauracion Neurologica, Havana, Cuba

## Appendix L Disclosures

- Dana Horakova received speaker honoraria and consulting fees from Biogen, Merck, Teva, Roche, Sanofi Genzyme, and Novartis, as well as support for research activities from Biogen and Czech Minsitry of Education [project Progres Q27/LF1].
- Eva Kubala Havrdova received honoraria/research support from Biogen, Merck Serono, Novars, Roche, and Teva; has been member of advisory boards for Actelion, Biogen, Celgene, Merck Serono, Novars, and Sanofi Genzyme; has been supported by the Czech Ministry of Educaon research project PROGRES Q27/LF1.
- Francesco Patti received speaker honoraria and advisory board fees from Almirall, Bayer, Biogen, Celgene, Merck, Novartis, Roche, Sanofi-Genzyme and TEVA. He received research funding from Biogen, Merck, FISM (Fondazione Italiana Sclerosi Multipla), Reload Onlus Association and University of Catania.
- Guillermo Izquierdo received speaking honoraria from Biogen, Novartis, Sanofi, Merck, Roche, Almirall and Teva.
- Sara Eichau received speaker honoraria and consultant fees from Biogen Idec, Novartis, Merck, Bayer, Sanofi Genzyme, Roche and Teva.
- Marc Girard received consulting fees from Teva Canada Innovation, Biogen, Novartis and Genzyme Sanofi; lecture payments from Teva Canada Innovation, Novartis and EMD. He has also received a research grant from Canadian Institutes of Health Research.
- Pierre Duquette served on editorial boards and has been supported to attend meetings by EMD, Biogen, Novartis, Genzyme, and TEVA Neuroscience. He holds grants from the CIHR and the MS Society of Canada and has received funding for investigator-initiated trials from Biogen, Novartis, and Genzyme.
- Alessandra Lugaresi has served as a Biogen, Bristol Myers Squibb, Merck Serono, Novartis, Roche, Sanofi/ Genzyme and Teva Advisory Board Member. She received congress and travel/accommodation expense compensations or speaker honoraria from Biogen, Merck, Mylan, Novartis, Roche, Sanofi/Genzyme, Teva and Fondazione Italiana Sclerosi Multipla (FISM). Her institutions received research grants from Novartis and Sanofi Genzyme.
- Pierre Grammond has served in advisory boards for Novartis, EMD Serono, Roche, Biogen idec, Sanofi Genzyme, Pendopharm and has received grant support from Genzyme and Roche, has received research grants for his institution from Biogen idec, Sanofi Genzyme, EMD Serono.
- Tomas Kalincik served on scientific advisory boards for BMS, Roche, Janssen, Sanofi Genzyme, Novartis, Merck and Biogen, steering committee for Brain Atrophy Initiative by Sanofi Genzyme, received conference travel support and/or speaker honoraria from WebMD Global, Eisai, Novartis, Biogen, Sanofi-Genzyme, Teva, BioCSL and Merck and received research or educational event support from Biogen, Novartis, Genzyme, Roche, Celgene and Merck.
- Raed Alroughani received honoraria as a speaker and for serving on scientific advisory boards from Bayer, Biogen, GSK, Merck, Novartis, Roche and Sanofi-Genzyme.
- Maria Pia Amato received honoraria as consultant on scientific advisory boards by Biogen, Bayer-Schering, Merck, Teva and Sanofi-Aventis; has received research grants by Biogen, Bayer-Schering, Merck, Teva and Novartis.
- Francois Grand’Maison received honoraria or research funding from Biogen, Genzyme, Novartis, Teva Neurosciences, Mitsubishi and ONO Pharmaceuticals.
- Katherine Buzzard received honoraria and consulting fees from Biogen, Teva, Novartis, Genzyme-Sanofi, Roche, Merck, CSL and Grifols.
- Murat Terzi received travel grants from Novartis, Bayer-Schering, Merck and Teva; has participated in clinical trials by Sanofi Aventis, Roche and Novartis.
- Cavit Boz received conference travel support from Biogen, Novartis, Bayer-Schering, Merck and Teva; has participated in clinical trials by Sanofi Aventis, Roche and Novartis.
- Jeannette Lechner-Scott travel compensation from Novartis, Biogen, Roche and Merck. Her institution receives the honoraria for talks and advisory board commitment as well as research grants from Biogen, Merck, Roche, TEVA and Novartis.
- Samia J. Khoury received compensation for participation in the Novartis Maestro program.
- Vincent van Pesch has received travel grants from Merck, Biogen, Sanofi, Bristol Myers Squibb, Almirall and Roche; his institution receives honoraria for consultancy and lectures and research grants from Roche, Biogen, Sanofi, Merck, Bristol Myers Squibb, Janssen, Almirall and Novartis Pharma.
- Radek Ampapa received conference travel support from Novartis, Teva, Biogen, Bayer and Merck and has participated in a clinical trials by Biogen, Novartis, Teva and Actelion.
- Julie Prevost accepted travel compensation from Novartis, Biogen, Genzyme, Teva, and speaking honoraria from Biogen, Novartis, Genzyme and Teva.
- Daniele Spitaleri received honoraria as a consultant on scientific advisory boards by Bayer-Schering, Novartis and Sanofi-Aventis and compensation for travel from Novartis, Biogen, Sanofi Aventis, Teva and Merck.
- Cristina Ramo-Tello received research funding, compensation for travel or speaker honoraria from Biogen, Novartis, Genzyme and Almirall.
- Claudio Solaro served on scientific advisory boards for Merck, Genzyme, Almirall, and Biogen; received honoraria and travel grants from Sanofi Aventis, Novartis, Biogen, Merck, Genzyme and Teva.
- Davide Maimone served on scientific advisory boards for Bayer, Biogen, Merck, Sanofi-Genzyme, Novartis, Roche, and Almirall; received honoraria and travel grants from Sanofi Genzyme, Novartis, Biogen, Merck, and Roche.
- Gerardo Iuliano (retired - no PI successor but has approved ongoing use of data) had travel/accommodations/meeting expenses funded by Bayer Schering, Biogen, Merck, Novartis, Sanofi Aventis, and Teva.
- Celia Oreja-Guevara received honoraria as consultant on scientific advisory boards from Biogen, Celgene, Merck, Novartis, Roche, Sanofi-Genzyme and TEVA.
- Bart Van Wijmeersch received research and rravel grants, honoraria for MS-Expert advisor and Speaker fees from Bayer-Schering, Biogen, Sanofi Genzyme, Merck, Novartis, Roche and Teva.
- Pamela McCombe received speakers fees and travel grants from Novartis, Biogen, T’évalua, Sanofi
- Mark Slee has participated in, but not received honoraria for, advisory board activity for Biogen, Merck, Bayer Schering, Sanofi Aventis and Novartis.
- Tamara Castillo-Triviño received speaking/consulting fees and/or travel funding from Bayer, Biogen, Merck, Novartis, Roche, Sanofi-Genzyme and Teva.
- Jose Luis Sanchez-Menoyo accepted travel compensation from Novartis, Merck and Biogen, speaking honoraria from Biogen, Novartis, Sanofi, Merck, Almirall, Bayer and Teva and has participated in clinical trials by Biogen, Merck and Roche
- Ricardo Fernandez Bolaños received speaking honoraria from Biogen, Novartis, Merck and Teva.
- Marie D’hooghe received consultancy and advisory board fees from Roche, Sanofi-Genzyme, Biogen, Merck-Serono, Bayer-Schering, Novartis and Allergan; received congress support from Biogen, Merck-Serono, Teva and Roche. She has also received research support from Novartis, Biogen, Roche, FWO (Research Foundation Flanders) and Fonds D.V. (Ligue Nationale Belge de la Sclerose en Plaques, Fondation Roi Baudouin).
- Guy Laureys received travel and/or consultancy compensation from Sanofi-Genzyme, Roche, Teva, Merck, Novartis, Celgene, Biogen.
- Anneke van der Walt served on advisory boards and receives unrestricted research grants from Novartis, Biogen, Merck and Roche She has received speaker’s honoraria and travel support from Novartis, Roche, and Merck. She receives grant support from the National Health and Medical Research Council of Australia and MS Research Australia.
- Helmut Butzkueven Institution (Monash university) has received compensation for consulting, talks, advisory / steering board activities from Biogen, Merck, Novartis, Genzyme, Alfred Health; research support from Novartis, Biogen, Roche, Merck, NHMRC, Pennycook Foundation, MSRA. HB has received compensation for same activities from Oxford Health Policy Forum, Merck, Biogen, Novartis.
- Michael Barnett served on scientific advisory boards for Biogen, Novartis and Genzyme and has received conference travel support from Biogen and Novartis. He serves on steering committees for trials conducted by Novartis. His institution has received research support from Biogen, Merck and Novartis.
- Jiwon Oh has received research funding from the MS Society of Canada, National MS Society, Brain Canada, Biogen, Roche, EMD Serono (an affiliate of Merck KGaA); and personal compensation for consulting or speaking from Alexion, Biogen, Celgene (BMS), EMD Serono (an affiliate of Merck KGaA), Novartis, Roche, and Sanofi-Genzyme.
- Ayse Altintas received speaker honoraria from Merck, Alexion,; received travel and registration grants from Merck, Biogen - Gen Pharma, Roche, Sanofi-Genzyme.
- Yara Fragoso received honoraria as a consultant on scientific advisory boards by Novartis, Teva, Roche and Sanofi-Aventis and compensation for travel from Novartis, Biogen, Sanofi Aventis, Teva, Roche and Merck.
- Edgardo Cristiano received honoraria as consultant on scientific advisory boards by Biogen, Bayer-Schering, Merck, Genzyme and Novartis; has participated in clinical trials/other research projects by Merck, Roche and Novartis.
- Tunde Csepany received speaker honoraria/ conference travel support from Bayer Schering, Biogen, Merck, Novartis, Roche, Sanofi-Aventis and Teva.
- Suzanne Hodgkinson received honoraria and consulting fees from Novartis, Bayer Schering and Sanofi, and travel grants from Novartis, Biogen Idec and Bayer Schering.
- Norma Deri received funding from Bayer, Merck, Biogen, Genzyme and Novartis.
- Shlomo Flechter received research funding, speaker honoraria and compensation for travel from and served as a consultant on advisory board for Bayer-Schering, Teva, Biogen, Merck, Genzyme and Novartis.
- Bruce Taylor received funding for travel and speaker honoraria from Bayer Schering Pharma, CSL Australia, Biogen and Novartis, and has served on advisory boards for Biogen, Novartis, Roche and CSL Australia.
- Fraser Moore participated in clinical trials sponsored by EMD Serono and Novartis.
- Orla Gray received honoraria as consultant on scientific advisory boards for Genzyme, Biogen, Merck, Roche and Novartis; has received travel grants from Biogen, Merck, Roche and Novartis; has participated in clinical trials by Biogen and Merck.
- Csilla Rozsa received speaker honoraria from Bayer Schering, Novartis and Biogen, congress and travel expense compensations from Biogen, Teva, Merck and Bayer Schering.
- Allan Kermode received speaker honoraria and scientific advisory board fees from Bayer, BioCSL, Biogen, Genzyme, Innate Immunotherapeutics, Merck, Novartis, Sanofi, Sanofi-Aventis, and Teva.
- Magdolna Simo received speaker honoraria from Novartis, Biogen, Bayer Schering; congress/travel compensation from Teva, Biogen, Merck, Bayer Schering.
- Cameron Shaw received travel assistance from Biogen and Novartis.
- Todd Hardy has received speaking fees or received honoraria for serving on advisory boards for Biogen, Merck, Teva, Novartis, Roche, Bristol-Myers Squibb and Sanofi-Genzyme, is Co-Editor of Advances in Clinical Neurosciences and Rehabilitation, and serves on the editorial board of Journal of Neuroimmunology and Frontiers in Neurology.
- Pamela McCombe received honoraria and consulting fees from Novartis, Bayer Schering and Sanofi and travel grants from Novartis, Biogen and Bayer Schering.
- Bhim Singhal received consultancy honoraria and compensation for travel from Biogen and Merck.
- Ilya Kister served on scientific advisory board for Biogen and received research support from Guthy-Jackson Charitable Foundation, National Multiple Sclerosis Society, Biogen,, and Novartis.
- Neil Shuey received travel compensation from Bayer Schering, Novartis, and Biogen Idec.
- Tatjana Petkovska-Boskova received congress and travel expense compensations from Biogen Idec and Teva.
- Nikolaos Grigoriadis received honoraria, consultancy/lecture fees, travel support and research grants from Biogen Idec, Biologix, Novartis, TEVA, Bayer, Merck Serono, Genesis Pharma, Sanofi – Genzyme, ROCHE, Cellgene, ELPEN and research grants from Hellenic Ministry of Development.

## Footnotes

1 The letter *w* was chosen because *worsening* is a shorthand for progression of disability. *w*_*c*_ stands for *confirmed* worsening.

